# Safety, Immunogenicity, and Efficacy of a COVID-19 Vaccine (NVX-CoV2373) Co-administered With Seasonal Influenza Vaccines

**DOI:** 10.1101/2021.06.09.21258556

**Authors:** Seth Toback, Eva Galiza, Catherine Cosgrove, James Galloway, Anna L. Goodman, Pauline A. Swift, Sankarasubramanian Rajaram, Alison Graves-Jones, Jonathan Edelman, Fiona Burns, Angela M. Minassian, Iksung Cho, Lakshmi Kumar, Joyce S. Plested, E. Joy Rivers, Andreana Robertson, Filip Dubovsky, Greg Glenn, Paul T. Heath, on behalf of the 2019nCoV-302 Study Group

## Abstract

**Background:** The safety and immunogenicity profile of COVID-19 vaccines when administered concomitantly with seasonal influenza vaccines has not yet been reported.

**Methods:** A sub-study on influenza vaccine co-administration was conducted as part of the phase 3 randomized trial of the safety and efficacy of NVX-CoV2373. The first ∼400 participants meeting main study entry criteria and with no contraindications to influenza vaccination were invited to join the sub-study. After randomization in a 1:1 ratio to receive NVX-CoV2373 (n=217) or placebo (n=214), sub-study participants received an age-appropriate, licensed, open-label influenza vaccine with dose 1 of NVX-CoV2373. Reactogenicity was evaluated via electronic diary for 7 days post-vaccination in addition to monitoring for unsolicited adverse events (AEs), medically-attended AEs (MAAEs), and serious AEs (SAEs). Influenza haemagglutination inhibition and SARS-CoV-2 anti-spike IgG assays were performed. Vaccine efficacy against PCR-confirmed, symptomatic COVID-19 was assessed. Comparisons were made between sub-study and main study participants.

**Findings:** Sub-study participants were younger, more racially diverse, and had fewer comorbid conditions than main study participants. Reactogenicity events more common in the co-administration group included tenderness (70.1% vs 57.6%) or pain (39.7% vs 29.3%) at injection site, fatigue (27.7% vs 19.4%), and muscle pain (28.3% vs 21.4%). Rates of unsolicited AEs, MAAEs, and SAEs were low and balanced between the two groups. Co-administration resulted in no change to influenza vaccine immune response, while a reduction in antibody responses to the NVX-CoV2373 vaccine was noted. Vaccine efficacy in the sub-study was 87.5% (95% CI: -0.2, 98.4) while efficacy in the main study was 89.8% (95% CI: 79.7, 95.5).

**Interpretation:** This is the first study to demonstrate the safety, immunogenicity, and efficacy profile of a COVID-19 vaccine when co-administered with seasonal influenza vaccines. The results suggest concomitant vaccination may be a viable immunisation strategy.

**Funding:** This study was funded by Novavax, Inc.

**Research in Context:** *Evidence before this study:* We searched PubMed for research articles published from December 2019 until 1 April 2021 with no language restrictions for the terms “SARS-CoV-2”, “COVID-19”, “vaccine”, “co-administration”, and “immunogenicity”. There were no peer-reviewed publications describing the simultaneous use of any SARS-CoV-2 vaccine and another vaccine. Several vaccine manufacturers had recent publications on phase 3 trials results (Pfizer/BioNTech, Moderna, AstraZeneca, Janssen, and the Gamaleya Research Institute of Epidemiology and Microbiology). Neither these publications nor their clinical trials’ protocols (when publicly available) described co-administration and they often had trial criteria specifically excluding those with recent or planned vaccination with any licenced vaccine near or at the time of any study injection.

*Added value of this study:* Immune interference and safety are always a concern when two vaccines are administered at the same time. This is the first study to demonstrate the safety and immunogenicity profile and clinical vaccine efficacy of a COVID-19 vaccine when co-administered with a seasonal influenza vaccine.

*Implications of all the available evidence:* This study provides much needed information to help guide national immunisation policy decision making on the critical issue of concomitant use of COVID-19 vaccines with influenza vaccines.

## INTRODUCTION

It is over a year since the start of the pandemic due to coronavirus disease 2019 (COVID-19) caused by the severe acute respiratory syndrome coronavirus 2 (SARS-CoV-2); a devastating disease with more than 170 million cases and 3.5 million deaths reported as of 2 June 2021.^1^ Seasonal influenza epidemics also occur globally and the World Health Organization (WHO) estimates that 290,000–650,000 individuals die from influenza each year, with the highest rates of death occurring in older adults and children younger than 2 years of age.^2^ Public health recommendations in many countries include yearly influenza vaccination as a key preventative strategy.^3^

Global COVID-19 vaccination efforts are now well underway with over 1.5 billion vaccine doses administered as of 31 May 2021.^1^ This continued mass COVID-19 vaccination programme will certainly coincide with influenza vaccination programmes. While the need for booster doses of COVID-19 vaccines has not yet been determined, the timing of such doses would likely overlap with the 2021–2022 influenza season in many settings. In addition, most countries will be still administering primary COVID-19 vaccine doses to the population when the need for influenza vaccines arises. Currently, there are no data regarding the co-administration of COVID-19 vaccines with other vaccines as most phase 3 trials of COVID-19 vaccines either excluded participants with recent or planned receipt of other licensed vaccines or required an interval of at least 1 week between them. In particular, knowledge of the effects of co-administration on immune responses and safety are needed to formulate public health policy in light of simultaneous vaccination programmes. This is particularly important as immunosenescence may leave older adults more vulnerable to influenza infection, complications, and mortality, as well as reduce their immune responses to standard influenza vaccines.^4^ Current guidance in the United Kingdom (UK) is to separate the administration of any deployed COVID-19 and influenza vaccines by at least 7 days to avoid incorrect attribution of potential adverse events (AEs).^3^ The Centers for Disease Control in the United States recommends a 14-day interval between these vaccines.^5^ However, the need for multiple clinic visits may lead to reduced compliance and hence reduced vaccination rates. To ensure adequate vaccine uptake of both COVID-19 and influenza vaccines, co-administration would encourage the public to take up these vaccines in one visit rather than returning 7 or more days later.

Herein we report the results of a sub-study of a phase 3 UK trial that assessed the safety and efficacy of two doses of NVX-CoV2373 compared with placebo.^6^ This sub-study aimed to evaluate the safety, immunogenicity, and efficacy of NVX-CoV2373 when co-administered with a licensed seasonal influenza vaccine.

## METHODS

### Trial Design and Participants

This influenza and COVID-19 vaccine co-administration study was a planned sub-study of a phase 3 UK trial that assessed the safety and efficacy of two 5-µg doses of NVX-CoV2373, administered intramuscularly 21 days apart, compared with placebo.^6^ Briefly, this study enrolled 15,139 participants at 33 sites in the UK beginning in September 2020. Eligible participants for the main trial were men and non-pregnant women 18 to 84 years of age (inclusive) who were healthy or had stable chronic medical conditions. Health status was assessed at screening and based on medical history, vital signs, and physical examination. Key exclusion criteria included a history of documented COVID-19, treatment with immunosuppressive therapy, or diagnosis with an unstable medical condition.

The first approximately 400 participants who met additional sub-study criteria were invited to participate in the influenza co-administration sub-study. Additional specific inclusion criteria included having not already received a 2020/2021 seasonal, licensed influenza vaccine and having no prior history of allergy or severe reaction to influenza vaccines. All participants were excluded from receipt of any live vaccine within 4 weeks or any vaccine within 2 weeks of the first dose of study vaccine or placebo co-administered with the influenza vaccine. Sub-study enrolment was not randomized or stratified by age.

All participants provided written informed consent before enrolment in the trial. The trial was designed and funded by Novavax. The trial protocol was approved by the North West—Greater Manchester Central Research Ethics Committee (Ref 20/NW/03/99) and was performed in accordance with the International Council for Harmonisation Good Clinical Practice guidelines.

Safety oversight was performed by an independent safety monitoring committee.

### Procedures

Seasonal influenza vaccine co-administration sub-study participants were selected prior to study vaccine randomisation. Approximately 400 consecutive, non-randomized, eligible participants from four study hospitals in the main study were enrolled. Participants were then randomly assigned in a 1:1 ratio via block randomization to receive two injections of NVX-CoV2373 or placebo (normal saline), 21 days apart. Randomization was stratified by site and by age ≥65 years. Participants in the seasonal influenza vaccine co-administration sub-study then received a concomitant dose of seasonal influenza vaccine with the first study injection only. This comprised a single intramuscular injection (0.5 mL) of a licensed influenza vaccine in the opposite deltoid to that of the study vaccine or placebo. Although the main study was observer-blinded, the influenza vaccine was administered in an open-label manner.

The study vaccine NVX-CoV2373 consisted of 5-μg SARS-CoV-2 rS with 50-μg Matrix-M adjuvant. Two influenza vaccines were utilised in the study to comply with Public Health England influenza vaccination recommendations^7^ (see Supplemental Material for influenza vaccine details).

- Influenza vaccine quadrivalent, cellular (QIVc) (Flucelvax^®^ Quadrivalent, Seqirus UK Limited, Maidenhead, UK) for those 18 to 64 years of age
- Adjuvanted trivalent influenza vaccine (aTIV) (Fluad^®^, Seqirus UK Limited, Maidenhead, UK) for those ≥65 years of age

### Immunogenicity Assessments

Blood was collected from all trial participants at baseline and at Day 21 for those in the influenza sub-study and for all trial participants at baseline and Day 35 (14 days after the second dose of study vaccine). A haemagglutination inhibition (HAI) assay antibody was performed in all influenza sub-study participants at baseline and at Day 21. An enzyme-linked immunosorbent assay (ELISA) for SARS-CoV-2 anti-Spike (anti-S) protein immunoglobulin G (IgG) was performed at baseline and on Day 35 in approximately 900 non-randomized participants from two study sites in the main study (as part of an immunogenicity cohort) as well as in those in the influenza sub-study (see Supplemental Material for additional assay details).

### Safety

After each study vaccination, participants remained under observation at the study site for at least 30 minutes to monitor for the presence of any acute reactions. Solicited local and systemic AEs were collected via an electronic diary for 7 days after each injection for approximately 2000 non-randomized participants from four study sites in the main study (as part of a reactogenicity cohort) as well as those in the influenza sub-study. Participants in the influenza sub-study were instructed to record local reactogenicity for the study vaccine (NVX-CoV2373 or placebo) injection site only. All participants were assessed for unsolicited AEs from the first injection or injections through 21 days; serious AEs (SAEs), AEs of special interest (AESIs) [including AESIs relevant to COVID and potentially-immune-mediated medical conditions (PIMMCs) – see Supplemental Tables S1 and S2)] and medically-attended AEs (MAAEs) were assessed from the first injection(s) through the end of the study period while only treatment-related MAAEs were analysed from the first injection(s) through Day 35. Unsolicited AEs and other safety events were reported for all participants who provided informed consent and received at least one injection in the main study and a co-administered influenza vaccine in the sub-study. Data from this ongoing phase 3 trial for the purpose of this analysis were assessed at a median of approximately 4 months after the first study injection (i.e. the dose with which influenza vaccine was co-administered). The safety follow-up period was the same for both the main study and sub-study. Participants in the influenza vaccine co-administration sub-study, the main study immunogenicity cohort, and main study reactogenicity cohort were all enrolled at separate, distinct locations.

### Efficacy

The primary efficacy endpoint was the first occurrence of virologically-confirmed symptomatic mild, moderate, or severe Covid-19, with onset at least 7 days after the second vaccination in participants who were seronegative at baseline. Symptomatic Covid-19 was defined according to US Food and Drug Administration (FDA) criteria.^6^ Symptoms of possible Covid-19 were assessed throughout the trial and collected using an electronic symptom diary for at least 10 days from symptom onset. At the onset of suspected Covid-19 symptoms, participants called their study site and when instructed, mucosal specimens from the nose and throat were collected daily over a 3-day period to assess for SARS-CoV-2 infection. Virological confirmation was performed using polymerase chain reaction testing. Daily temperature self-measurements were recorded at home for at least 10 days and participants were evaluated for an initial clinical assessment (in 1–3 days). A follow-up assessment was conducted (in 7–10 days) where physical examinations were performed and vital signs were collected.

### Statistical Analysis

#### Safety Analysis

Unsolicited AEs, SAEs, MAAEs, and AESIs were analysed in all participants who received at least one dose of NVX-CoV2373 or placebo for the main study and one dose of NVX-CoV2373 or placebo plus one dose of influenza vaccine for the sub-study. Safety events were summarised descriptively. Solicited local and systemic AEs after the first injection(s) were also summarised by FDA toxicity grading criteria and duration after each injection (see **Supplementary Table S3**). Unsolicited AEs were coded by preferred term and system organ class using Version 23.1 of the *Medical Dictionary for Regulatory Activities* (MedDRA) and summarised by severity and relationship to study vaccine. Participants in the sub-study were then compared with participants in the main study, by study vaccine and influenza vaccine received (NVX-CoV2373 plus influenza vaccine; NVX-CoV2373 alone; placebo plus influenza vaccine; placebo alone).

### Immunogenicity Analysis

For participants who received the influenza vaccine, strain-specific immune responses to influenza vaccine as measured by HAI and reported as geometric mean titres (GMTs), geometric man fold-rise (GMFR) comparing at Day 0 (baseline) and at Day 21, and seroconversion rates (SCRs) (defined as the proportion of subjects with either a baseline reciprocal titre of <10 and a post-vaccination reciprocal titre ≥40, or a baseline titre of ≥10 and a post-vaccination titre ≥4-fold higher). For influenza strain-specific GMTs according to group (influenza vaccine concomitantly administered with NVX-CoV2373 or with placebo), titres reported below the lower limit of quantitation (LLOQ; i.e. below the starting dilution of assay reported as “<10”) were set to half that limit (i.e. 10 / 2 = 5).

For the SARS-CoV-2 anti-S protein IgG antibody levels measured by the ELISA assay, geometric mean ELISA units (GMEUs) at each study visit (Day 0 and Day 35), the geometric mean fold rises (GMFRs) comparing at Day 0 and at Day 35, along with 95% CI, were summarised by vaccine group (NVX-CoV2373 plus influenza vaccine; NVX-CoV2373 alone; placebo plus influenza vaccine; placebo alone). Data were also assessed by age group (18 to <65, ≥65 to 84) and corresponding influenza vaccine types (QIVc and aTIV, respectively). The SCR for the IgG antibody was defined as a proportion of participants with ≥4-fold rises. ELISA units (EUs) reported below the lower limit of quantitation (LLOQ; i.e. below the starting dilution of assay reported as “<200”) were set to half that limit (i.e. 200 / 2 = 100).

For both HAI and IgG antibody measured by treatment group, the 95% CIs were calculated based on the t distribution of the log-transformed values, then back transformed to the original scale for presentation as GMTs/GMEUs and GMFRs. The SCRs, along with 95% CIs based on the Clopper-Pearson method, were summarised by vaccine group. The per-protocol immunogenicity analysis set was defined as those who received two doses of vaccine, had all immunology samples available, had no major protocol deviations, and did not have a laboratory confirmed SARS-CoV-2 infection prior to any visit where serology was measured.

Non-randomized comparisons of the Day 35 anti-S EUs were performed using a geometric mean ratio (GMR) defined as the ratio of two GMEUs. An analysis of covariance on log transformed values with group, age, and baseline EUs was performed. The ratios of geometric least square means and 95% CIs for the ratios were calculated by back transforming the mean differences and 95% confidence limits for the differences of log (base 10) transformed EUs between the two groups. The two-sided 95% CIs for the absolute rate difference between two groups were constructed using the Newcombe method.

### Efficacy Analysis

The trial was designed and driven by the total number of events expected to achieve statistical significance for the primary endpoint – a target of 100 mild, moderate, or severe Covid-19 cases for the main study. The target number of 100 cases for the final analysis provides >95% power for 70% or higher vaccine efficacy. The main (hypothesis testing) event-driven analysis for the final analyses of the primary objective was carried out at an overall one-sided type I error rate of 0.025 for the primary endpoint. The primary endpoint (per-protocol population) was analysed in participants who were seronegative at baseline, received both doses of study vaccine or placebo, had no major protocol deviations affecting the primary endpoint, and had no confirmed cases of symptomatic Covid-19 from the first dose until 6 days after the second dose (per-protocol efficacy population). Vaccine efficacy was defined as VE (%) = (1 – RR) × 100, where RR = relative risk of incidence rates between the two study groups (NVX-CoV2373 or placebo). The estimated RR and its CI for the main study were derived using Poisson regression with robust error variance.^8^ Hypothesis testing of the primary endpoint was carried out against the null hypothesis: H0: vaccine efficacy ≤30%. The study met success criterion by rejecting of the null hypothesis to demonstrate a statistically significant vaccine efficacy.

## RESULTS

### Participants

Between 28 September and 28 November 2020, a total of 15,187 participants were randomised into the main phase 3 trial of which 431 were co-vaccinated with a seasonal influenza vaccine (QIVc or aTIV, depending on participant age); 217 sub-study participants received NVX-CoV2373 and 214 received placebo. In the influenza sub-study group, 43.3 % were female, 75.1% were White, 22.7% were from ethnic minorities or reported multiple races, 27.1% had at least one comorbid condition (based on Centers for Disease Control and Prevention definitions^5^). The median age of sub-study participants was 39 years, 32.9% were 50 years of age or older, and 6.7 % were 65 years of age or older (see **Supplementary Table S4**). Within the sub-study, there were 29 aTIV recipients with a median age of 66 years (n=16 in the NVX-CoV2373 arm) and 69 years (n=13 in the placebo arm) and 402 QIVc recipients with a median age of 38 years (n=201 in the NVX-CoV2372 arm) and 37 years (n=201 in the placebo arm) (**Table 1**). A total of 431 participants were assessed for unsolicited AEs, SAEs, MAAEs, and AESIs, while 404 participated in the assessment of reactogenicity. All 431 participants were part of the evaluable immunogenicity population for both HAI and anti-S IgG assays. The sub-study group overall was younger, more racially diverse, and had fewer comorbid conditions than participants in the main study and the main study reactogenicity and immunogenicity cohorts (**Table 1**, **Supplementary Tables S4 and S5**). The main study immunogenicity cohort for the anti-S IgG assay included 999 participants in the intention-to-treat population who had received either the NVX-CoV2373 vaccine or placebo alone. The main study reactogenicity cohort included 2310 from the safety population who had received at least one dose of the NVX-CoV2373 vaccine or placebo alone.

**Table 1:** Demographics and baseline characteristics of participants in the influenza vaccine co-administration sub-study and entire study populations (ITT population)

|  | **NVX-CoV2373**<br>**+ aTIV**<br>**(n=16)** | **NVX-CoV2373**<br>**+ QIVc**<br>**(n=201)** | **Placebo**<br>**+ aTIV**<br>**(n=13)** | **Placebo**<br>**+ QIVc**<br>**(n=201)** | **Total Study, ITT**<br>**Population**<br>**(n=15139)** |
| --- | --- | --- | --- | --- | --- |
| Age, yr (SD) | 66.9 (1.86) | 40.3 (12.72) | 69.3 (3.73) | 40.2 (11.57) | 53.1 (14.91) |
| Median | 66.0 | 38.0 | 69.0 | 37.0 | 55.0 |
| Range | 65, 71 | 20, 64 | 65, 77 | 23, 64 | 18, 84 |
| Age group, n (%) |  |  |  |  |  |
| 18-64 yr | 0 (0) | 201 (100) | 0 (0) | 201 (100) | 11014 (72.8) |
| ≥65 yr | 16 (100) | 0 (0) | 13 (100) | 0 (0) | 4125 (27.2) |
| Sex, n (%) |  |  |  |  |  |
| Male | 6 (37.5) | 117 (58.2) | 4 (30.8) | 114 (56.7) | 7808 (51.6) |
| Female | 10 (62.5) | 84 (41.8) | 9 (69.2) | 87 (43.3) | 7331 (48.4) |
| Race or ethnic group, n (%) |  |  |  |  |  |
| White | 12 (75.0) | 151 (75.1) | 11 (84.6) | 153 (76.1) | 14280 (94.3) |
| Black or African American | 0 (0) | 4 (2.0) | 0 | 2 (1.0) | 60 (0.4) |
| Asian | 0 (0) | 14 (7.0) | 1 (7.7) | 22 (10.9) | 462 (3.1) |
| Multiple | 4 (25.0) | 25 (12.4) | 0 (0) | 23 (11.4) | 136 (0.9) |
| Not reported | 0 (0) | 3 (1.5) | 1 (7.7) | 1 (0.5) | 176 (1.2) |
| Other | 0 (0) | 3 (1.5) | 0 (0) | 0 (0) | 17 (<0.1) |
| Missing | 0 (0) | 1 (0.5) | 0 (0) | 0 (0) | 8 |
| Hispanic or Latinx | 1 (6.3) | 9 (4.5) | 1 (7.7) | 4 (2.0) | 125 (0.8) |
| quadrivalent |  |  |  |  |  |
| SARS-CoV-2 serostatus, n (%) |  |  |  |  |  |
| Negative | 15 (93.8) | 183 (91.0) | 12 (92.3) | 184 (91.5) | 14362 (94.9) |
| Positive | 1 (6.3) | 18 (9.0) | 0 (0.0) | 13 (6.5) | 643 (4.2) |
| Missing | 0 (0) | 0 (0) | 1 (0.7) | 4 (2.0) | 134 (0.9) |
| Comorbidity status* |  |  |  |  |  |
| Yes | 5 (31.3) | 50 (24.9) | 7 (53.8) | 55 (27.4) | 6767 (44.7) |
| No | 11 (68.8) | 151 (75.1) | 6 (46.2) | 146 (72.6) | 8372 (55.3) |

\*Comorbid subjects are those identified who have at least one of the comorbid conditions reported as a medical history or have a screening body mass index value greater than 30 kg/m<sup>2</sup>.
Percentages are based on the intention-to-treat data set within the seasonal influenza vaccine sub-study (by vaccine type; aTIV for those ≥65 years of age and QIVc for those <65 years of age) and overall.
Abbreviations: aTIV=adjuvanted trivalent influenza vaccine; ITT=intention-to-treat; QIVc=influenza vaccine quadrivalent, cellular; SD=standard deviation.

### Safety and Reactogenicity

Overall local reactogenicity (assessed only at the non-influenza vaccine injection site) was largely absent or mild in the co-administration group, NVX-CoV2373 alone group, and placebo plus influenza vaccine group (**Figure 1**). Any local reaction was reported in 70.1% of those co-vaccinated (1.7% severe), 57.6% in the NVX-CoV2373 alone group (1.0% severe), 39.4% (0% severe) in the placebo plus influenza vaccine group, and 17.9% (0.2% severe) in the placebo alone group. The most commonly reported local reactions were injection site tenderness and injection site pain, occurring in 64.9% and 39.7% of those co-vaccinated and 53.3% and 29.3% of those given NVX-CoV2373 alone, respectively.

**Figure 1:**
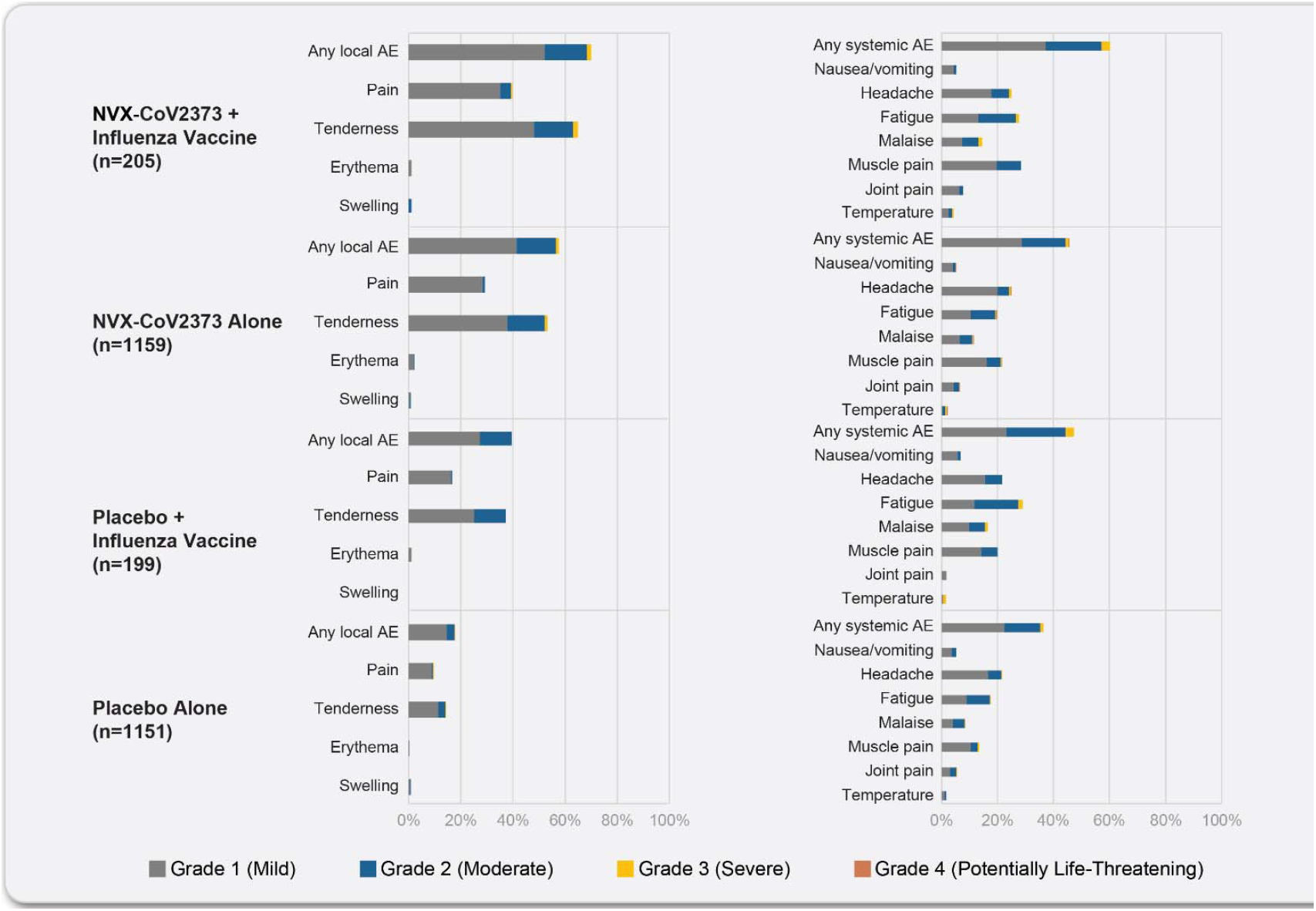
Reactogenicity data from participants in the influenza vaccine co-administration sub-study and participants in the reactogenicity cohort population after dose 1: local and systemic. The percentage of participants in each treatment group with solicited local and systemic adverse events during the 7 days after each vaccination is plotted according to the maximum toxicity grade (mild, moderate, severe, or potentially life-threatening) in participants included in the seasonal influenza vaccine sub-study and those included in the reactogenicity cohort.

Any systemic reaction was reported in 60.1% of those co-vaccinated (2.9% severe), 45.7% in the NVX-CoV2373 alone group (1.3% severe), 47.2% in the placebo plus influenza vaccine group (2.8% severe), and 36.3% (1.1% severe) in the placebo alone group. In general, the incidence of specific systemic reactogenicity events was similar within all of these groups (**Figure 1**). The most commonly reported systemic events were muscle pain and fatigue, occurring in 28.3% and 27.7% of those co-vaccinated and 21.4% and 19.4% of those given NVX-CoV2373 alone, respectively, with muscle pain (28.3%) also occurring more frequently in the co-administration group than the placebo plus influenza vaccine group (20.0%). Notably, fever (temperature ≥38°C) was reported in 4.3%, 2.0%, 1.7%, and 1.5% in the co-vaccinated, NVX-CoV2373 alone, placebo plus influenza vaccine, and placebo alone groups, respectively (see **Supplementary Tables S6–S9**).

When assessed by specific influenza vaccine type, QIVc in those <65 years of age and aTIV in those ≥65 years of age, among those administered concomitantly with NVX-CoV2373, there was a trend towards lower rates of local and systemic reactogenicity in the older group who received the aTIV. Of note, the median duration of reactogenicity events was generally 1–2 days for local events and approximately 1 day for systemic events in both the co-vaccinated group and the NVX-CoV2373 alone group. When assessed by specific influenza vaccine type, there was a general trend for a shorter duration of reactogenicity among those ≥65 years of age (aTIV recipients) (data not shown).

Unsolicited AEs reported up to 21 days after first vaccination were predominantly mild in severity and were similarly distributed across the co-vaccinated and NVX-CoV2373 alone groups (**Table 2**). The frequency of all and severe AEs in the co-vaccinated group (18.4% and 0.5%, respectively) was similar to those in the NVX-CoV2373 alone group (17.6% and 0.4%, respectively). These rates were also similar to the rates of all and severe AEs in the placebo plus influenza vaccine group (14.5% and 0.0%, respectively) and placebo alone group (14.0% and 0.4%, respectively). The unsolicited AEs occurring in >1% of the co-vaccinated group included headache (2.3%), fatigue (1.8%), and oropharyngeal pain (1.4%). Rates of all MAAEs were 7.8% and 3.8% in those co-vaccinated and those who received NVX-CoV2373 alone, respectively, while rates of MAAEs in the placebo plus influenza vaccine group and placebo group alone were 8.4% and 3.9%, respectively. Rates of treatment-related MAAEs were lower and balanced in all groups (**Table 2**). The rate of SAEs was also low and balanced among the sub-study participants and those not involved in the sub-study. No treatment related SAEs were reported in sub-study participants. No PIMMCs and/or AESIs relevant to COVID-19 were seen in the influenza co-administration sub-study, with resulting event rates similar to those not involved in the sub-study. There were no episodes of anaphylaxis or deaths within the sub-study.

**Table 2:** Safety data from participants in the influenza vaccine co-administration sub-study and participants in the entire study population (without sub-study participants)

|  | **NVX-CoV2373 +**<br>**Influenza Vaccine** | **Placebo +**<br>**Influenza Vaccine** | **NVX-CoV2372 Alone** | **Placebo Alone** |
| --- | --- | --- | --- | --- |
|  | n=217 | n=214 | n=7352 | n=7356 |
| Any AE | 40 (18.4%) | 31 (14.5%) | 1297 (17.6%) | 1030 (14.0%) |
| Any severe AE | 1 (0.5%) | 0 (0%) | 33 (0.4%) | 33 (0.4%) |
| SAE | 1 (0.5%) | 0 (0%) | 43 (0.6%) | 44 (0.6%) |
| MAAE | 17 (7.8%) | 18 (8.4%) | 279 (3.8%) | 288 (3.9%) |
| Treatment-related MAAE | 3 (1.4%) | 0 (0%) | 34 (0.5%) | 17 (0.2%) |
| PIMMC | 0 (0%) | 0 (0%) | 5 (<0.1%) | 8 (0.1%) |
| AESI related to COVID | 0 (0%) | 0 (0%) | 8 (0.1%) | 22 (0.3%) |

Influenza vaccine co-administration sub-study participants compared with the entire ITT study population, excluding the co-vaccination sub-study group. Adverse events and severe adverse events are those within 21 days of study dose 1 (with or without co-administration of influenza vaccine). SAEs, MAAEs, AESIs, and PIMMCs are assessed for the entire study period.
Abbreviations: AE=adverse event; AESI=adverse event of special interest; ITT, intention-to-treat; MAAE=medically-attend adverse event; PIMMC= potentially-immune-mediated medical condition; SAE=serious adverse event.

### Immunogenicity

#### Response to influenza vaccine

There were no statistically significant differences in baseline HAI GMT titres between those in the sub-study co-vaccinated with NVX-CoV2373 plus influenza vaccine group and those in the placebo plus influenza vaccine group (**Figure 2A&B**). In the QIVc groups, HAI GMTs were significantly higher after vaccination on Day 21 while in the much smaller aTIV groups, there was overlap in GMT CIs before and after vaccination. No difference in Day 21 HAI GMTs was seen between the NVX-CoV2373 plus influenza vaccine group and the placebo plus influenza vaccine group for any individual influenza strain (A/H1N1, A/H3N2, B/Victoria, or B/Yamagata) for either influenza vaccine. GMFR values followed the same pattern (see specific strain information in **Supplementary Table S10** and **Table S11**). For both QIVc and aTIV, HAI SCRs were generally high for the influenza A strains but lower for the influenza B strains (**Figure 3A&B**).

**Figure 2:**
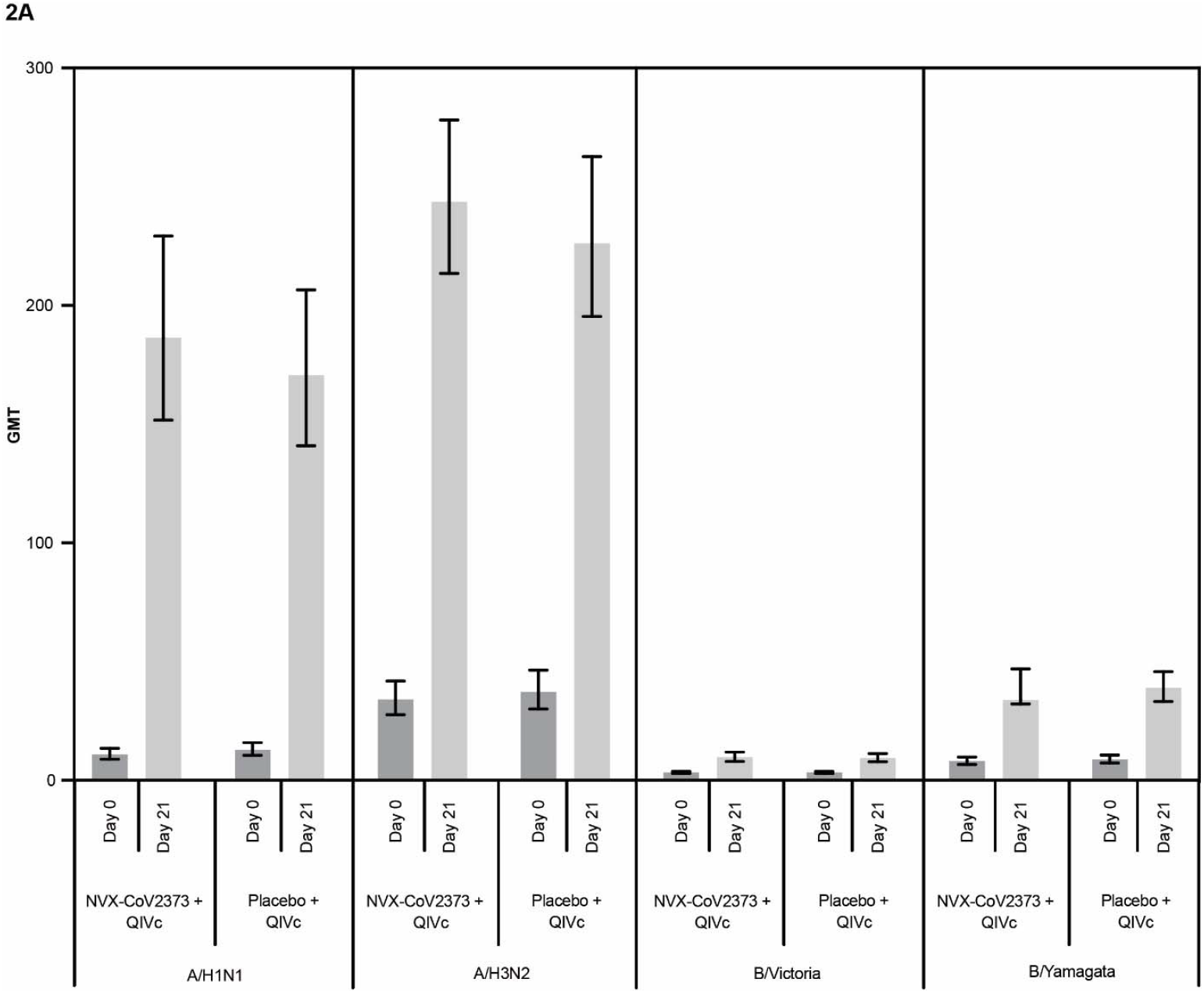
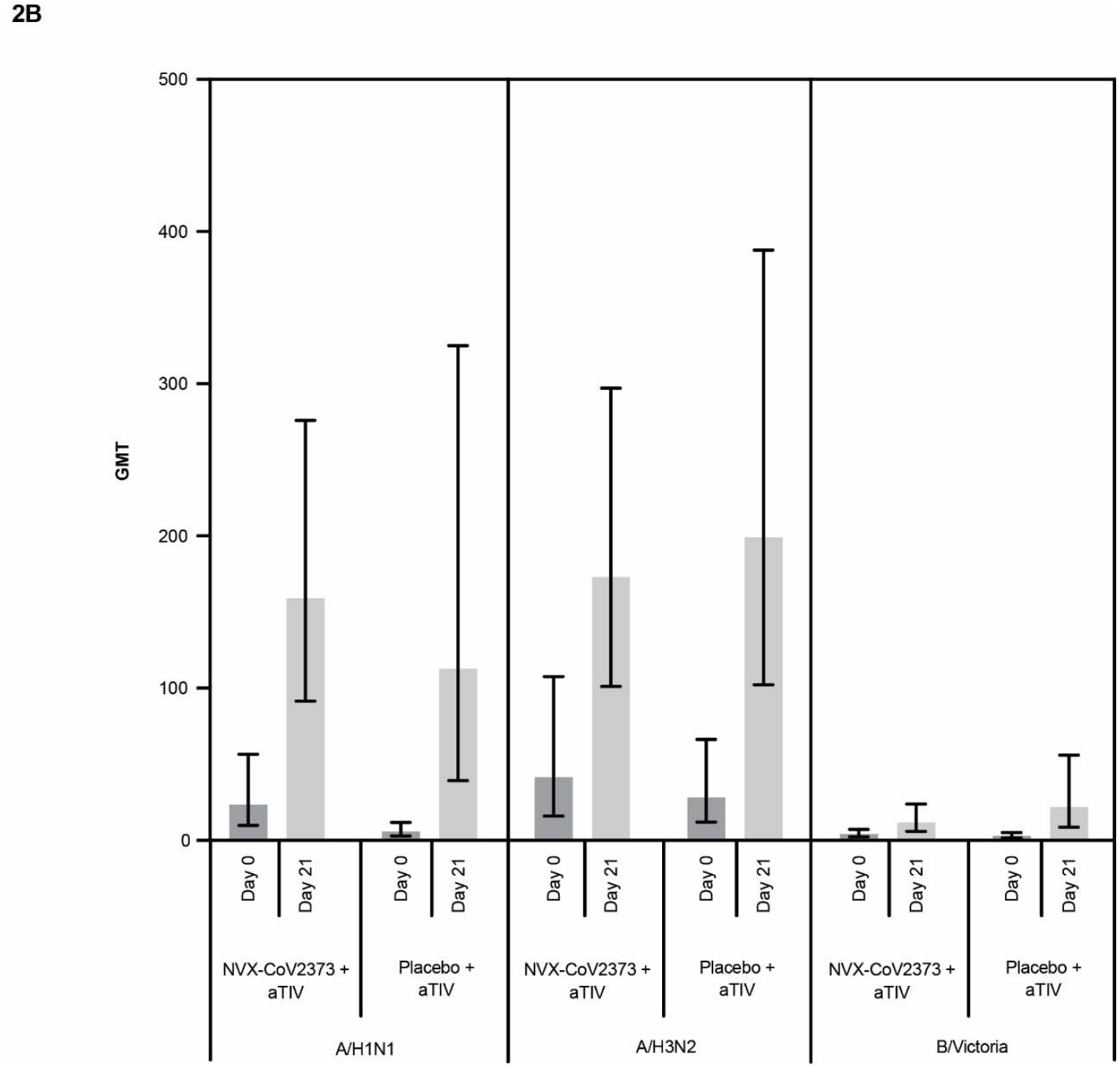
A) HAI GMTs on Day 0 and Day 21 in the aTIV Group; B) HAI GMTs on Day 0 and Day 21 in the QIVc Group. Comparison of the HAI GMTs at baseline (Day 0) and 21 days after vaccination with NVX-CoV2373 or placebo with either aTIV or QIVc influenza vaccine by influenza strain. aTIV= adjuvanted trivalent influenza vaccine; GMT=geometric mean titre; HAI=haemagglutination inhibition; QIVc=influenza vaccine quadrivalent, cellular.

**Figure 3:**
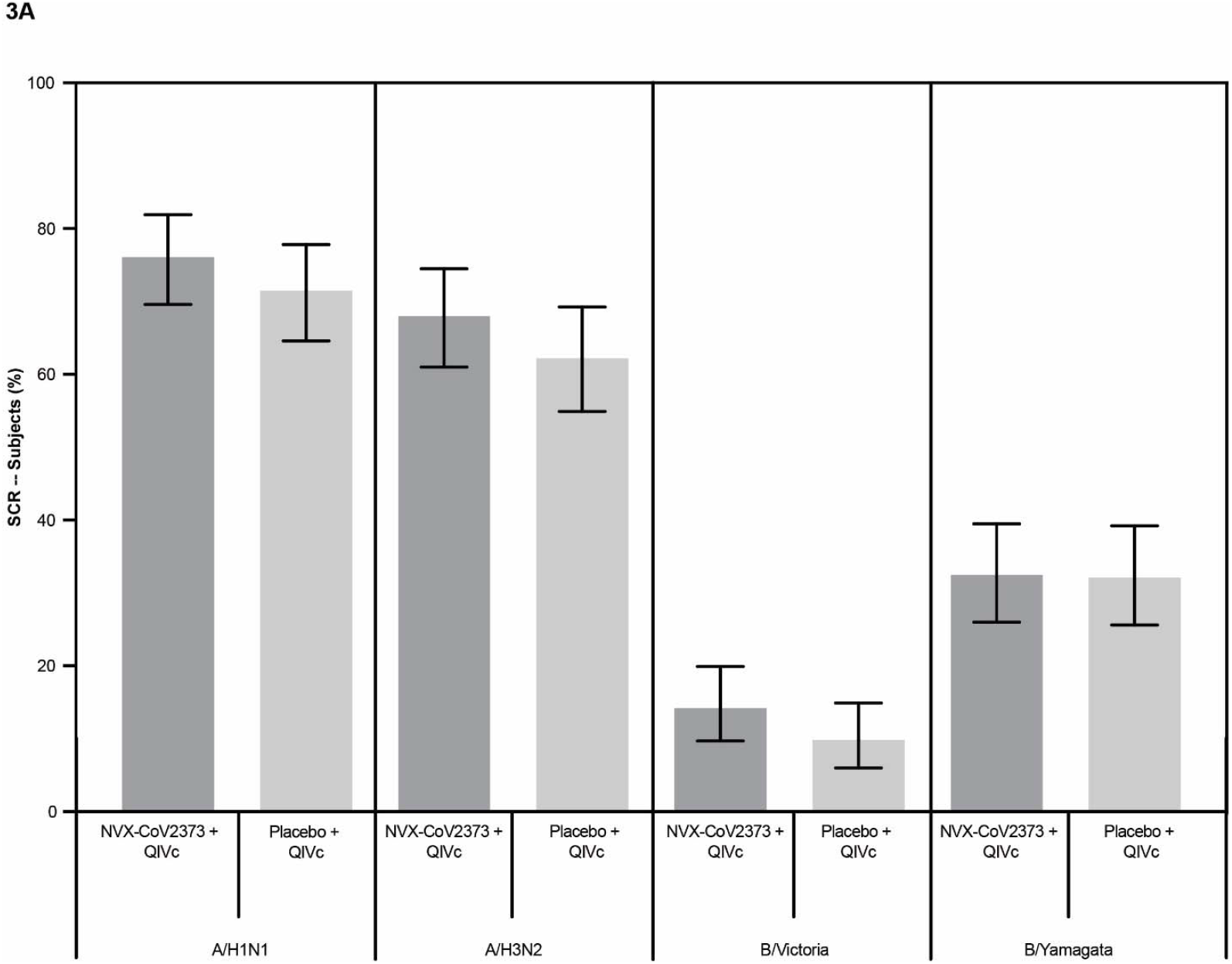
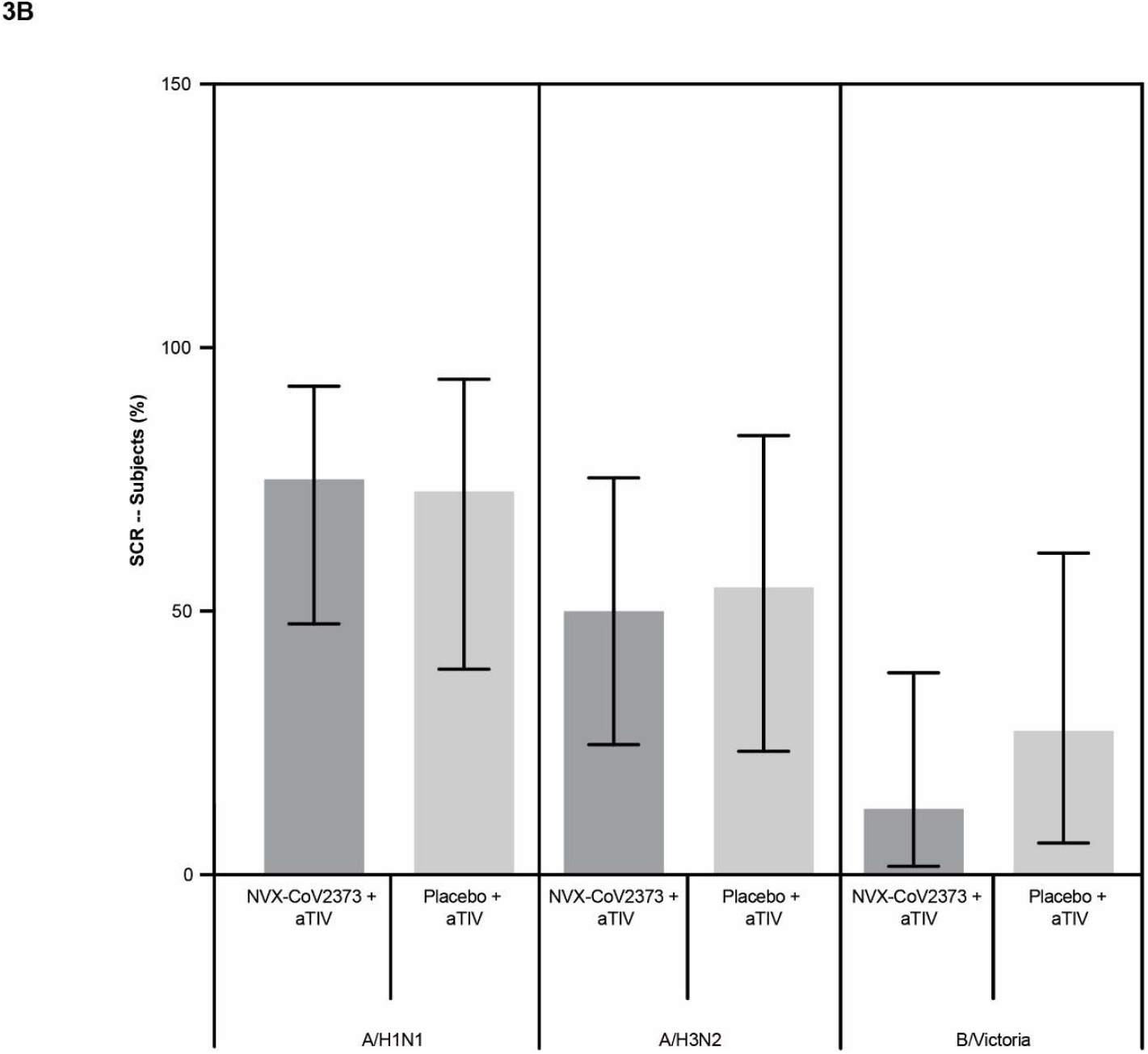
A) HAI SCRs on Day 0 and Day 21 in the aTIV Group; B) HAI SCRs on Day 0 and Day 21 in the QIVc Group. Comparison of the HAI SCRs 21 days after vaccination with NVX-CoV2373 or placebo with either aTIV or QIVc influenza vaccine by influenza strain. aTIV= adjuvanted trivalent influenza vaccine; HAI=haemagglutination inhibition; QIVc=influenza vaccine quadrivalent, cellular; SCR=seroconversion rate.

#### Response to NVX-CoV2373

Baseline anti-S EUs were similar in participants in the sub-study co-vaccinated with NVX-CoV2373 and influenza vaccine and those who received placebo plus influenza vaccine as well as in those vaccinated in the main study immunogenicity cohort with NVX-CoV2373 alone (data for the immunogenicity per-protocol population are in **Table 3**. In both groups vaccinated with NVX-CoV2373 plus influenza vaccine or with NVX-CoV2373 alone, the Day 35 GMEUs were significantly higher than those at baseline. A difference in GMEUs was observed between the two per-protocol groups (NVX-CoV2373 plus influenza vaccine [n=178]: 31,236.1 [95% CI: 26,295.51, 37,104.9] vs. NVX-CoV2373 alone [n=414]: 46,678.3 [95% CI: 40,352.2, 49,468.2]). A post hoc assessment of the ratio between the two geometric means when adjusted for baseline EUs, age, and treatment group was 0.57 (95% CI: 0.47, 0.70). This difference was also reflected in the GMFRs, but not in the SCRs, which were 97.8% and 99.0% in the two groups, respectively. The Day 35 GMEUs were numerically lower in the ≥65-year-old (aTIV) concomitant vaccination group compared with the 18- to <65-year-old (QIVc) concomitant vaccination group, although the number of participants in the concomitant aTIV group was small. However, the GMFRs were large, >200, and the SCRs were both >97%. This diminution in immunogenicity with increasing age was also seen in the main study immunogenicity cohort. The subgroup of participants receiving concomitant NVX-CoV2373 and any influenza vaccine who were seropositive (n=19) at baseline achieved Day 35 GMEUs that were significantly greater than those in similar participants who were seronegative (n=198) at baseline (71,115.6 [95% CI: 46,813.8, 108,032.8] vs. 30,439.1 [95% CI: 25,713.4, 36,033.5], respectively) (see **Supplementary Table S12A**).

**Table 3:**
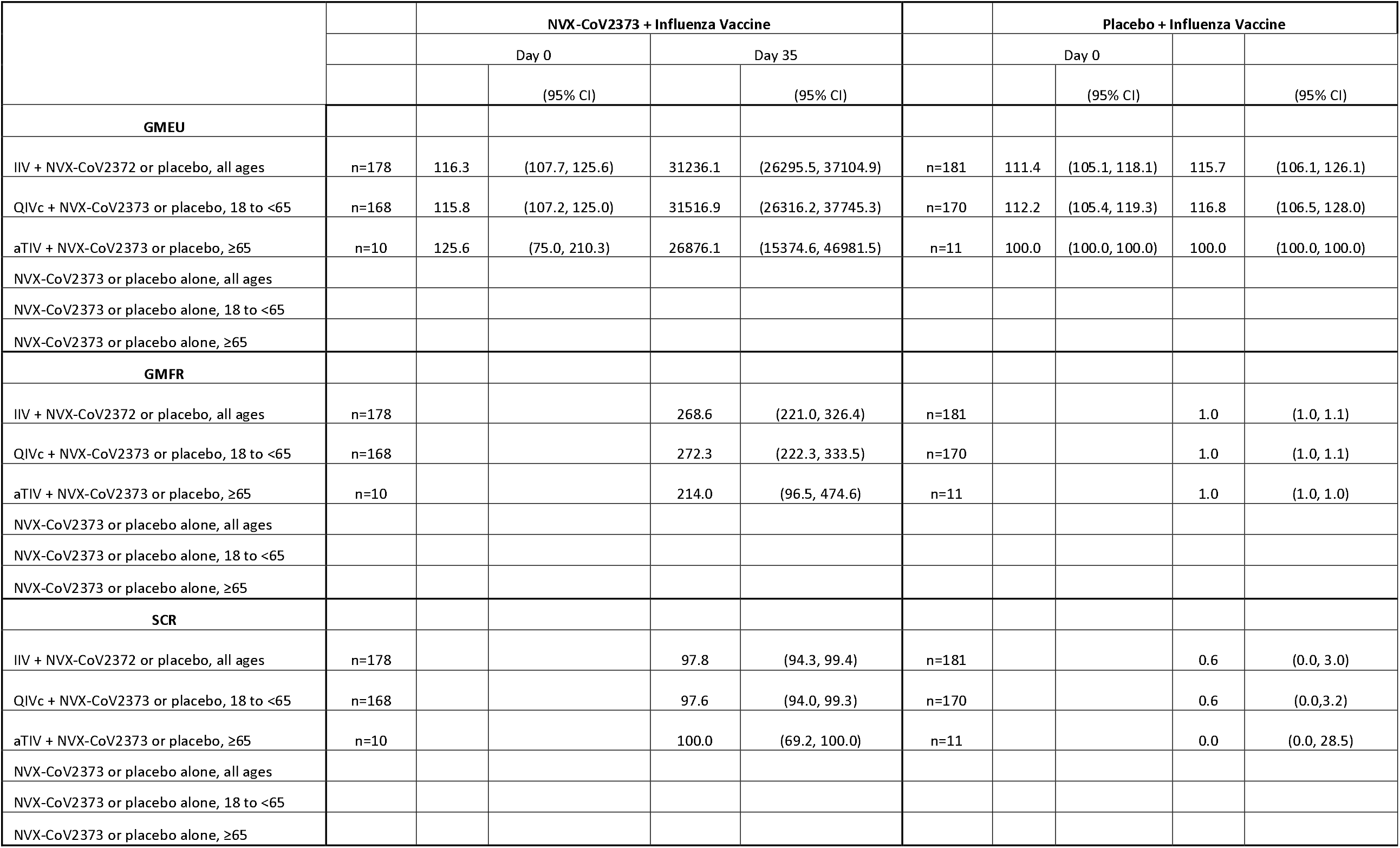
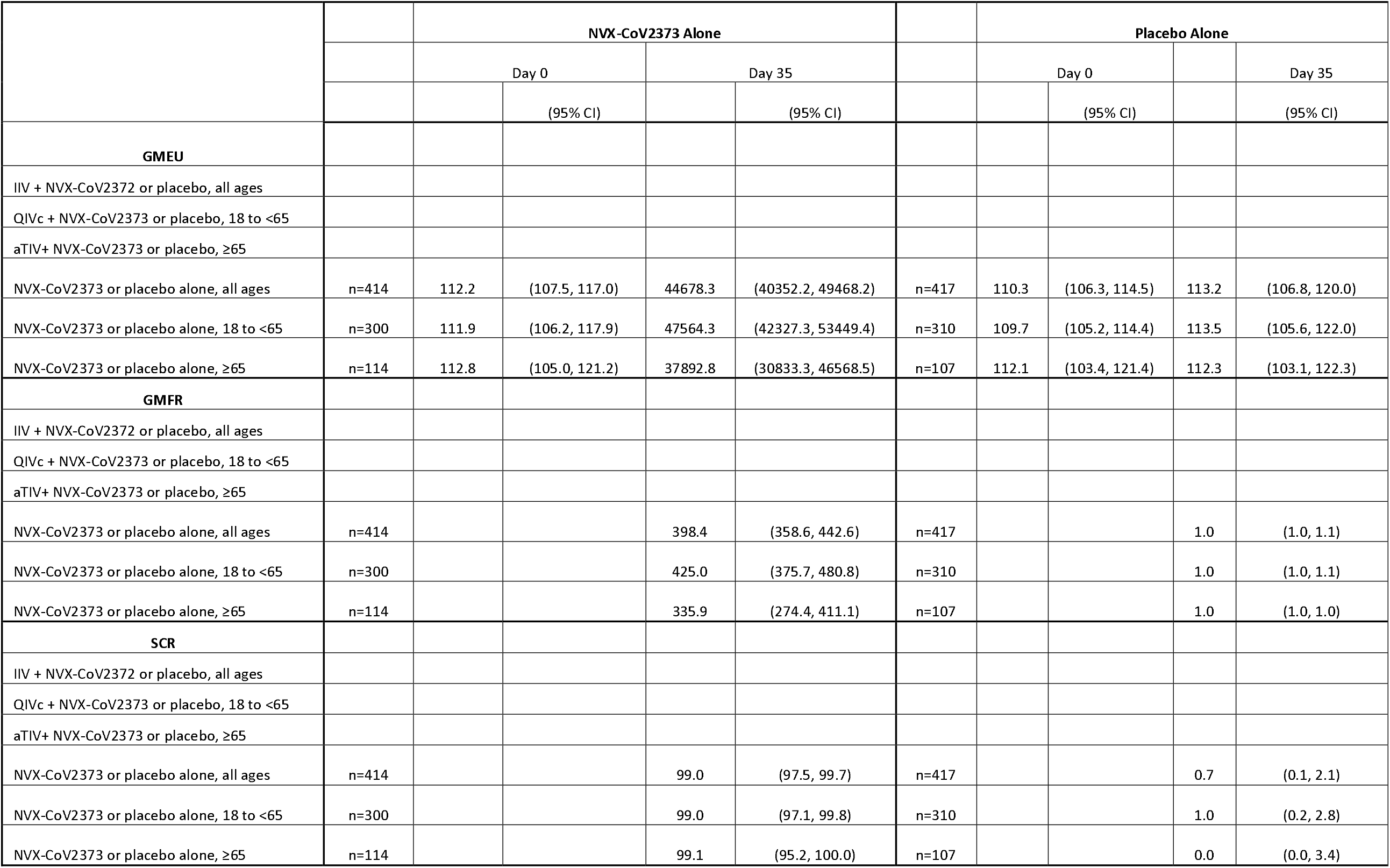
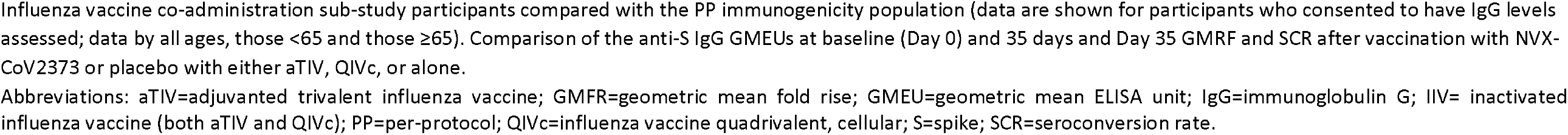
Anti-S IgG on Day 0 and Day 35 in the influenza vaccination sub-study and immunogenicity cohort, in the PP population, by age group.

### Efficacy

Among 360 participants in the influenza sub-study who were also in the efficacy per-protocol population who were 18 to <65 years of age, there was one case of virologically-confirmed, symptomatic Covid-19 with onset at least 7 days after the second dose among vaccine recipients and eight cases among placebo recipients. A post hoc analysis of the primary endpoint among those 18 to <65 years of age demonstrated a vaccine efficacy of 87.5% (95% CI, -0.2 to 98.4) (**Table 4**). There were too few cases among those in the per-protocol population who were ≥65 years to calculate a vaccine efficacy. All influenza sub-study cases among those 18 to <65 years of age were due to the B.1.1.7 variant. Vaccine efficacy in the entire per-protocol population 18 to <65 years of age was of 89.8% (95% CI, 79.7 to 95.5) while vaccine efficacy against the B.1.1.7 variant alone in per-protocol population was of 86.3% (95% CI, 71.3 to 93.5).

**Table 4:** Vaccine efficacy against PCR-confirmed symptomatic Covid-19 with an onset at least 7 days after second study vaccination in serologically negative participants.

| Parameter | Analysis |  |
| --- | --- | --- |
|  | NVX-CoV2373 | Placebo |
| Participants, 18 to <65 years, per-protocol influenza sub-study, n | 178 | 182 |
| Participants with first occurrence of event, n | 1 | 8 |
| Vaccine efficacy (%) * | 87.5 |  |
| 95% CI | -0.2, 98.4 |  |
| Participants, 18 to <65 years, per-protocol population, n | 5067 | 5062 |
| Participants with first occurrence of event, n | 9 | 87 |
| Vaccine efficacy (%) | 89.8 |  |
| 95% CI | 79.7, 95.5 |  |
| Participants, per-protocol population due to B.1.1.7 variant, n | 7020 | 7020 |
| Participants with first occurrence of event, n | 8 | 58 |
| Vaccine efficacy (%) | 86.3 |  |
| 95% CI | 71.3, 93.5 |  |

\*All strains were characterized as B.1.1.7 variants. Vaccine efficacy as assessed in those 18 to <65 years of age in the influenza co-administration sub-study within the per-protocol population. Vaccine efficacy was also calculated for the same population within the main study and in all per-protocol participants infected with the B.1.1.7 variant.
Abbreviations: CI=confidence interval; PCR=polymerase chain reaction.

## DISCUSSION

This study is the first to demonstrate the safety, immunogenicity, and efficacy of any COVID-19 vaccine when co-administered with a seasonal influenza vaccine or any other vaccination. Most COVID-19 vaccine trials have excluded participants receiving other vaccinations at the time or near the time of injection with study vaccine and therefore have no interaction studies addressed in their labels^9–11^ Although no specific comparative immunogenicity endpoints were pre-specified in this exploratory sub-study, we found no evidence for interference of the COVID-19 vaccine with the QIVc influenza vaccine. Definitive conclusions about aTIV were not possible because of the small number of participants older than 65 years of age. We did, however, observe an impact of concomitant administration of an influenza vaccine on the absolute magnitude of the anti-S antibody response. This impact did not seem to be clinically meaningful as vaccine efficacy appeared to be preserved. Co-administration also appeared to have no clinically meaningful effect on systemic or local reactogenicity and no additional safety concerns were found to be associated with co-vaccination. Solicited local and systemic reactogenicity events after co-administration were generally similar to the incidence and severity of those for each vaccine when administered separately. The incidence of more subjective local reactogenicity (pain and tenderness) was elevated in the co-vaccinated group above the level of either the NVX-CoV2373 alone or placebo plus influenza vaccine groups, but the rates for more objective local events (erythema and swelling) were low and indistinguishable between all groups. These increased rates were largely driven by an increase in mild symptoms. It is unclear if subjects were biased in their assessment of pain and tenderness at the study injection site having received two co-administered vaccinations; the fact that placebo injections were assessed as causing more local pain/tenderness when given concomitantly with an influenza vaccine (in the opposite arm) compared with placebo injections, when given alone, would suggest this is likely to be the case. Another explanation is that participants recorded local symptoms from the influenza injection site despite being instructed to consider the injection site of the study vaccine only. The rate for any systemic reactogenicity event was modestly elevated over the rate for either NVX-CoV2373 or influenza vaccine alone, consistent with an overall higher vaccine immunogen load and the relatively younger participant population in the sub-study. This was seen mainly for the events of muscle pain and fever, yet despite the relative increase in the rate of fever, the absolute fever rate was modest (4.3%). Rates of severe events were low in all groups and showed no clinically meaningful pattern of increased reactogenicity. The elevation in some reactogenicity events may, in part, have been due to the overall younger age of the influenza vaccine sub-study participants compared with the main study reactogenicity cohort (median age 39.0 years [93.3% 18 to <65 years] vs. a median age of 52.0 years [80.1% 18 to <65 years]). Those ≥65 years of age who received two adjuvanted vaccines compared with those <65 years of age who received the adjuvanted NVX-CoV2373 and unadjuvanted QIVc had lower rates of reactogenicity; this effect of age was also seen in the NVX-CoV2373 alone group and in prior NVX-CoV2373 studies^6,12,13^ and is consistent with immunosenescence.

The rates of AEs, SAEs, and AESIs were low and balanced between those given NVX-CoV2373, influenza vaccine, or both. The rate of any MAAE was higher in sub-study participants compared with non-sub-study participants. This difference was less apparent when assessing treatment-related MAAEs only. The increased rate of all MAAEs in the sub-study may represent a health-care seeking bias in those desiring an influenza vaccine rather than a true increase in medical visits due to AEs related to co-vaccination or receipt of the influenza vaccine plus placebo; an assessment of these excess medical visits revealed that most were general practice visits associated with health maintenance concerns (data not shown).

The magnitude of the humoral response to either influenza vaccine was not affected by co-administration with NVX-CoV2372 when assessed at 21 days after dosing, although care should be used in generalising this observation to aTIV because of the small sample size. The post-vaccination rise in GMTs and SCRs for each strain were high when either influenza vaccine was administered with placebo or NVX-CoV2373, although there was a generally lower response to the influenza B strains found in all influenza vaccine recipients. The humoral immune response to influenza B strains is dependent upon numerous factors including age and prior influenza vaccine exposure.^14^ Low influenza B SCRs^15^ and lower SCRs relative to influenza A strains^16,17^ have been seen with prior immunogenicity studies of quadrivalent inactivated influenza vaccines.

In contrast, there was a modest reduction in the anti-S EUs observed with the co-administration of NVX-CoV2373 and an influenza vaccine. It is unclear if this reduction was due to vaccine interference or due to the non-randomized nature of the studied groups. In the absence of a correlate of protection, it is difficult to interpret the significance of this finding. The post hoc assessment of vaccine efficacy in this sub-study in those 18 to <65 years of age was 87.5% compared with the vaccine efficacy of 89.8% in the same age group from the per-protocol efficacy populations in the main study; although given the small number of endpoint cases in the sub-study the lower bound of the confidence interval was just below zero. The similar vaccine efficacy within the influenza vaccine co-administration group would suggest that the reduction in the anti-S EUs as a result of co-administration may not be clinically meaningful. In fact, the levels of anti-S EUs in those receiving both vaccines (in either those 18 to <65 or ≥65 years of age) was still over 3-fold greater than the anti-S EUs found in convalescent serum, suggesting that EUs in this range found in sub-study participants may be protective.^18^ It should be also noted that no difference in the rates of SCRs were seen between those co-vaccinated and those who received NVX-CoV2373 alone.

It is also apparent that the extent of the reduction in anti-S EUs may be less relevant in participants who are seropositive at baseline, as they achieved high values post-vaccination with co-administration of influenza vaccine with a mean of 71,115 EUs in co-vaccinated seropositive participants of all ages compared with a mean of 44,678 EUs in per-protocol NVX-CoV2373 alone recipients of all ages (**Table 3** and **Supplementary Table S12A**). One possible explanation for this finding is that seropositive individuals have pre-existing T-cell and B-cell populations with immune memory against the SARS-CoV2 spike protein minimizing any possible effect of immune interference. Therefore, it is possible that influenza vaccine co-administration may impact priming but have no impact on the immune response in previously primed individuals. This observation could inform public health vaccination strategy such as suggesting that influenza vaccine co-administration occur with the second dose of any two-dose COVID-19 vaccine (as concomitant administration was only performed with the first dose in the current study). The same mechanism in curtailing potential immune interference may become apparent if influenza vaccines are given concomitantly with COVID-19 vaccine booster doses.

Although this is the first study to show the co-administration of a COVID-19 with a seasonal influenza vaccine, influenza vaccine co-administration has been well studied. Our study utilised two different influenza vaccines for different age groups in compliance with UK influenza vaccination guidelines.^19^ For those <65 years of age, a cell culture–derived, inactivated quadrivalent influenza vaccine was used. QIVc was approved in the UK in December 2018 for individuals 9 years and older and extended to 2 years and older in 2020. For the older cohort, a MF59 squalene-based, oil-in-water aTIV was administered. This aTIV was approved in the UK in August 2017. In two studies of the MF59 aTIV given concomitantly with a pneumococcal vaccine, antibody responses to either vaccine were not affected and the safety data were consistent with expected rates of AEs for both vaccines.^20,21^ No interference or safety concerns have been reported with a QIV co-administered with pneumococcal and herpes zoster vaccines.^22,23^

The strengths of this sub-study include the placebo-controlled design and its alignment with national influenza vaccine policy in the use of both adjuvanted and unadjuvanted influenza vaccines in different age groups. Study limitations include the small overall sub-study size (with few participants ≥65 years of age owing to the high rate of routine influenza vaccination among participants in this age group at study start), small number of sub-study efficacy endpoints, lack of formal pre-specified non-inferiority statistical assessment of immunogenicity, and the lack of randomization in recruiting the influenza sub-study, immunogenicity, and reactogenicity cohorts. A stronger design could have been four randomized arms consisting of NVX-CoV2373 plus influenza vaccine, NVX-CoV2373 plus placebo, influenza vaccine plus placebo, and placebo plus placebo. Another limitation was the open-label design in administering the influenza vaccine, but this was required to order to allow participants to consider only the study vaccine injection site for assessment of local symptoms. Finally, the assessment of neutralising antibody titres may have benefitted the immunogenicity investigation, yet prior studies with NVX-CoV2373 have shown a strong correlation between the anti-S and wild-type microneutralizations results.^18^

This is the first study to demonstrate the safety, immunogenicity, and efficacy profile of a COVID-19 vaccine when co-administered with a seasonal influenza vaccine. These data demonstrate no early safety concerns with the concomitant administration of NVX-CoV2373 with an influenza vaccine. Immunogenicity of the influenza vaccine was preserved with concomitant administration while a modest decrease in the immunogenicity of the NVX-CoV2373 vaccine was found. Vaccine efficacy in those 18 to <65 years appeared to be preserved in those receiving both vaccines compared with those vaccinated with NVX-CoV2373 alone. Future clinical trials and post-licensure studies of COVID-19 vaccines should include safety and immunogenicity data on co-administration with common adult and paediatric vaccines. More research on the concomitant vaccination of COVID-19 and influenza vaccines is needed, especially in those >65 years of age, to help guide national immunisation policy on this critical issue.

## Data Availability

The protocol for this phase 3 study is publicly available from Novavax.

https://www.clinicaltrialsregister.eu/ctr-search/trial/2020-004123-16/GB

## Contributors

ST, JSP, LK, FD, GG, IC, AR, and EJR are Novavax employees. PTH is the chief investigator. ST, PTH, JP, LK, FD, GG, IC, and AR contributed to the protocol and design of the study. EG, CG, ALG, JG, FB, AMM and PAS are study site principal investigators. SR, JE, and AG are Seqirus employees. EG, CG, ALG, JG, FB, AMM and PS contributed to the study or data collection. IC and AR conducted the statistical analysis. All authors reviewed, commented on, and approved this manuscript prior to submission for publication.

## Data sharing

The protocol for this phase 3 study is publicly available from Novavax.

## Acknowledgements

The study and article were funded by Novavax. We would like to thank all the study participants for their commitment to this study. We also acknowledge the investigators and their study teams for their hard work and dedication. In addition, we would like to thank the National Institute for Health Research, representatives from the Department of Health and Social Care laboratories, and NHS Digital and the members of the UK Vaccine Task Force. Editorial assistance in the preparation of this manuscript was provided by Phase Five Communications, funded by Novavax, Inc.

