## Supplemental Materials for "Safety, Immunogenicity, and Efficacy of a COVID-19 Vaccine (NVX-CoV2373) Co-administered With Seasonal Influenza Vaccines"

This supplementary material has been provided by the authors to give readers additional information about their work.

Table S2. Adverse Events of Special Interest Relevant to COVID-19 ………………………………………….	10

Table S3. FDA Toxicity Grading Scale for Clinical Abnormalities

(Local and General Systemic Reactogenicity) …………………………………………………………………...	11

Table S4. Demographics for the Influenza Vaccination Sub-Study Population, Reactogenicity Cohort,

and Intention-to-Treat Group (Without the Influenza Sub-Study Participants) ………………………………….	12

Table S5. Demographics for the Anti-S IgG Immunogenicity Cohort and Intention to Treat Population ………	13

Table S6. Reactogenicity for Participants in the Influenza Vaccination Sub-Study and Reactogenicity

Cohort After Dose 1, All Ages …………………………………………………………………………………...	14

Table S7. Reactogenicity for Participants in the Influenza Vaccination Sub-Study and Reactogenicity

Cohort After Dose 1, Participants 18 to <65 Years Old …………………………………………………………	15

Table S8. Reactogenicity for Participants in the Influenza Vaccination Sub-Study and Reactogenicity

Cohort After Dose 1, Participants ≥65 Years Old ……………………………………………………………….	16

Table S9. Reactogenicity for Participants in the Influenza Vaccination Sub-Study After Dose 2

(No Concomitant Influenza Vaccine), All Ages …………………………………………………………………	17

Table S10. Influenza Haemagglutination Inhibition Assay: GMT, GMFR, and SCR Values by

Influenza Strain for QIVc …………………………………………………………………………………………	18

Table S11. Influenza Haemagglutination Inhibition Assay: GMT, GMFR, and SCR Values by

Influenza Strain for aTIV …………………………………………………………………………………………	19

Table S12a&b. Anti-S IgG on Day 0 and Day 35 in the Influenza Vaccination Sub-Study (a) and Immunogenicity

Cohort (b) by Serostatus, in the ITT Population, All Ages ….…………………………………………………...	20

**2019nCoV-302 Study Group Members**

The NVX-CoV2373-2019nCoV-302 clinical trial was a collective group effort across multiple institutions and locations. Below is a list of sites and staff that significantly contributed to the implementation and conduct of the NVX-CoV2373-2019nCoV-302 clinical trial.

| **Site** | **Investigators** |
| --- | --- |
| Aberdeen Royal Infirmary, NHS Grampian | Roy L. Soiza, Robin Brittain-Long, Chiara Scicluna, Carole Edwards, Lynn Mackay, Mariella D’Allesandro, Amy Nicol, Karen Norris, Sandra Mann, Heather Lawrence, Ruth Valentine |
| Accelerated Enrollment Solutions | Marianne Elizabeth Viljoen, Carol H. Pretswell, Helen Nicholls, Imrozia Munsoor, Agnieszka Meyrick, Christina Kyriakidou, Shalini Iyengar, Arham Jamal, Nick Richards, Helen Price, Bridie Rowbotham, Danielle Bird, Karen Smith, Olga Littler, Kirsty Fielding, Anna Townsend-Rose, Karen Miller, Jessica Davis, Alison Elliot-Garwood, Lauren Trottier, Paul Edwards |
| Belfast Health and Trust | Margaret McFarland |
| Betsi Cadwaladr University Health Board | Orod Osanlou, Laura Longshaw, Jane Stockport, Victoria Saul, Alice Thomas, Lynne Grundy |
| Blandford Group Practice | Katharine Lucy Broad |
| Bradford Teaching Hospitals NHS Foundation Trust | Karen Regan, Kim Storton, Declan Ryan-Wakeling, Brad Wilson, Malathy Munisamy, John Wright, Anil Shenoy, Beverley English, Lucy Brear |
| Centre for Clinical Vaccinology and Tropical Medicine, University of Oxford | Paola Cicconi |
| Chelsea and Westminster Hospital | Marta Boffito, Ana Milinkovic, Ruth Byrne, Roya Movahedi, Rosalie Housman |
| County Durham & Darlington NHS Foundation Trust | Naveed Kara, Ellen Brown |
| Department of Psychiatry, University of Oxford, NIHR Oxford Health Cognitive Health Clinical Research Facility and NIHR Oxford Health Biomedical Research Centre | Andrea Cipriani, Mary-Jane Attenburrow, Katharine A. Smith |
| Division of Epidemiology and Public Health, University of Nottingham | Jonathan Packham |
| Dorset Research Hub, Royal Bournemouth Hospital, University Hospitals Dorset NHS Foundation Trust | Geoff Sparrow |
| East Suffolk and North Essex NHS Foundation Trust | Richard Smith, Josephine M Rosier, Khalid Saja, Nyasha Nago, Brian Camilleri, Anita Immanuel, Mike Hamblin, Rawlings Osagie, Mahalakshmi Mohan |
| Epsom and St Helier University Hospitals NHS Trust | Hilary Floyd, Suzanne Goddard, Sanjay Mutgi, John Evans, Sean McKeon, Neringa Vilimiene, Rosavic Chicano, Rachel Hayre, Alice Pandaan |
| Faculty of Health and Life Sciences, Oxford Brookes University | Catherine Henshall |
| Health and Care Research Wales | Nicola Williams, Jayne Goodwin |
| Highcliffe Medical Centre | Zelda Cheng |
| Keele University | Toby Helliwell, Adrian Chudyk |
| Kings College London | Rafaela Giemza, John Lord Villajin, Noah Yogo, Esther Makanju, Pearl Dulawan, Deepak Nagra, April Buazon, Alice Russell, Georgie Bird |
| Lakeside Healthcare Research, Lakeside Surgery | Amardeep Heer, Rex Sarmiento, Balraj Sanghera, Melanie Mullin, Adam Champion, Aisling Bevan, Kinzah Iqbal, Alshia Johnson |
| Layton Medical Centre | Rebecca Clark, Sarah Shaw, Steven Shaw, Amanda Chalk, Martin Lovatt, Caroline Lillicrap, Angela Parker, Jan Hansel, Zhi Wong, Galvin Gan, Eyad Tuma |
| Leeds Teaching Hospitals NHS Trust | Jane Minton, Jennifer Murira, Razan Saman, Alistair Hall, Kyra Holliday, Zara Khan, James Calderwood, George Twigg, Helena Baker, Julie Corrigan, Katy Houseman |
| Midlands Partnership NHS Foundation Trust | Subhra Raguvanshi, Dominic Heining, Jake Weddell, Liz Glaves, Kim Thompson, Francis Davies, Ruth Lambley Burke |
| MRC-University of Glasgow Centre for Virus Research, and Queen Elizabeth University Hospital, NHS Greater Glasgow & Clyde, Glasgow, Scotland | Emma C. Thomson |
| Guy's and St Thomas' NHS Foundation Trust NIHR BRC | Sonia Serrano, Andrea Mazzella, Thurkka Rajeswaran, Moncy Mathew, Karen Bisnauthsing, Laura Bremner, Henry Fok, Franca Morselli, Paola Cinardo, Blair Merrick, Lucy Sowole |
| National Institute for Health Research Patient Recruitment Centre and Bradford Teaching Hospitals NHS Foundation Trust, Bradford | Dinesh Saralaya |
| NIHR Clinical Research Facility, University Hospital Southampton NHS Foundation Trust | Lisa Berry |
| NIHR Southampton Clinical Research Facility and NIHR Wessex Local Clinical Research Network, University Hospital Southampton NHS Foundation Trust | Mihaela Pacurar, Saul N Faust |
| NIHR Clinical Research Network, Thames Valley and South Midlands, Oxford University Hospitals NHS Foundation Trust | Nancy Hopewell, Leigh Gerdes |
| Norfolk and Norwich University Hospital NHS Foundation Trust | Jeremy Turner, Christopher Jeanes, Adele Cooper, Jocelyn Keshet-Price, Lou Coke, Melissa Cambell-Kelly, Ketan Dhatariya, Claire Williams , Georgina Marks, James Sudbury, Lisa Rodolico |
| Northern Ireland Clinical Research Facility, Queen's University Belfast and Belfast Health and Social Care Trust | Judy Bradley, Sharon Carr, Roisin Martin, Angelina Madden, Paul Biagioni, Sonia McKenna, Alison Clinton |
| Northern Ireland Clinical Research Network | Maurice O’Kane |
| North Tees and Hartlepool NHS Foundation Trust | Justin Carter, Matthew Dewhurst, Bill Wetherill |
| Oxford Health NHS Foundation Trust, Warneford Hospital | Thandiwe Hoggarth, Katrina Lennon Collins, Marie Chowdhury, Adil Nathoo, Anna Heinen, Orla MacDonald, Claudia Hurducas, Liliana Cifuentes, Harjeevan Gill, Andy Gibson, Raha West |
| Quadram Institute | Jane Ewing |
| Queen Elizabeth University Hospital, NHS Greater Glasgow and Clyde | Guy Mollett, Rachel Blacow, John Haughney, Jonathan MacDonald, John Paul Seenan, Stewart Webb, Colin O'Leary, Scott Muir, Beth White, Neil Ritchie |
| Queen's University Belfast and Belfast Health and Social Care Trust | Daniel F. McAuley, Jonathan Stewart |
| Research and Development, NHS Grampian | Chiara Scicluna, Mariella D'Alessandro, Carole Edwards, Lynn MacKay, Amy Nicol, Karen Norris, Heather Lawrence, Sandra Mann, Ruth Valentine |
| Royal Bournemouth Hospital, University Hospitals Dorset NHS Foundation Trust | Nicki Lakeman, Laura Purandare |
| Royal Cornwall Hospital NHS Trust | Duncan Browne, David Tucker, Peter Luck, Angharad Everden, Lisa Trembath, Michael Visick, Nick Morley, Laura Reid, Helen Chenoweth, Kirsty Maclean |
| Royal Devon and Exeter Hospital | Ray P. Sheridan, Tom Burden, Craig Francis Lunt, Shirley Todd, Stephanie Estcourt, Jasmine Marie Pearce, Suzanne Wilkins, Cathryn Love-Rouse |
| Royal Free London NHS Foundation Trust | Eva Torok-Pollok, Mike Youle, Sara Madge, Danielle Solomon, Melissa Chowdhury, Aarti Nandani, Janet North, Nargis Hemat, Suluma Mohamed |
| Royal Oldham Hospital, Northern Care Alliance, Greater Manchester | Rachel Newport |
| Salford Royal Hospital, Northern Care Alliance, Greater Manchester | Philip A. Kalra, Chukwuma Chukwu, Olivia Wickens, Vikki O'Loughlin, Hema Mistry, Louise Harrison, Robert Oliver, Anne-Marie Peers, Jess Zadik, Katie Doyle |
| South Tees Hospitals NHS Foundation Trust | David R. Chadwick, Kerry Colling, Caroline Wroe, Marie Branch, Alison Chilvers, Sarah Essex |
| Stafford Town Primary Care Network | Mark Stone |
| Vaccine Institute, St George’s University of London & St George's University Hospitals NHS Foundation Trust | Alberto San Francisco Ramos, Emily Beales, Olivia Bird, Zsofia Danos, Hazel Fofie, Cecilia Hultin, Sabina Ikram, Fran Mabesa, Aoife Mescall, Josyanne Pereira, Jennifer Pearce, Natalina Sutton |
| St Helens and Knowsley Teaching Hospitals NHS Trust | Emma Snashall |
| Stockport NHS Foundation Trust, Stepping Hill Hospital | David Neil Baxter, Sara Bennett, Debbie Suggitt, Kerry Hughes, Wiesia Woodyatt, Lynsey Beacon, Alissa Kent, Chris Cooper, Milan Rudic, Simon Tunstall, Matthew Jackson |
| Swanage Medical Practice | Claire Hombersley |
| The Adam Practice | Patrick Moore, Rebecca Cutts |
| University College London | Danielle Solomon, Janet M. North |
| University Hospitals of Morecambe Bay NHS Foundation Trust | Andrew Higham, Marwan Bukhari, Mohamed Elnaggar, Michelle Glover, Fiona Richardson, Alexandra Dent, Shahzeb Mirza, Rajiv Ark, Jennie Han |
| University of Exeter Medical School, William Wright House, Royal Devon and Exeter Hospital | Suzy V Hope, Philip J Mitchelmore |
| University of Liverpool | Rostam Osanlou, Thomas Heseltine |

**Supplemental Methods**

*Details on Vaccines*

The influenza vaccine quadrivalent (QIVc) (Flucelvax^®^ Quadrivalent, Seqirus UK Limited, Maidenhead, UK) is indicated for those 18 to 64 years of age. The QIVc dose contains 15 µg haemagglutinin (HA) per 0.5 mL dose from each of the following strains: A/Hawaii/70/2019 (H1N1)pdm09-like strain (A/Nebraska/14/2019, wild type), A/Hong Kong/45/2019 (H3N2)-like strain (A/Delaware/39/2019, wild type), B/Washington/02/2019-like strain (B/Darwin/7/2019, wild type), and B/Phuket/3073/2013-like strain (B/Singapore/INFTT-16-0610/2016, wild type).

The adjuvanted trivalent influenza vaccine (aTIV) (Fluad^®^, Seqirus UK Limited, Maidenhead, UK) is indicated for those ≥65 years of age. Within each 0.5 mL dose, the aTIV formulation contains the MF59 adjuvant (9.75 mg squalene, 1.175 mg polysorbate 80, 1.175 mg sorbitan trioleate, 0.66 mg sodium citrate, 0.04 mg citric acid, water for injections) plus 15 µg HA from each of the following strains: A/Guandong-Maonan/SWL1536/2019 (H1N1)pdm09-like strain (A/Victoria/2454/2019 IVR-207), A/Hong Kong/2671/2019 (H3N2)-like strain (A/Hong Kong/2671/2019 IVR-208), and B/Washington/02/2019-like strain (B/Victoria/705/2018 BVR-11).

Strains assessed in the haemagglutination inhibition assay (HAI):

*Quadrivalent formulation of cell- or recombinant-based influenza vaccine*

- A/Nebraska/14/2019 (an A/Hawaii/70/2019 (H1N1)pdm09-like virus)
- A/Delaware/39/2019 (an A/Hong Kong/45/2019 (H3N2)-like virus)
- B/Darwin/7/2019 (a B/Washington/02/2019-like virus)
- B/Singapore/INFTT-16-0610/2016 (a B/Phuket/3073/2013-like virus)

*Trivalent formulation of egg-based influenza vaccine*

- A/Victoria/2454/2019 IVR-207 (an A/Guangdong-Maonan/SWL1536/2019 (H1N1) pdm09-like virus
- A/Hong Kong/2671/2019 IVR-208 (an A/Hong Kong/2671/2019 (H3N2)-like virus)
- B/Victoria/705/2018 BVR-11 (a B/Washington/02/2019-like virus).

*Details on the Assays*

*HAI*

The haemagglutination inhibition assay was performed at the Public Health England Laboratory, Porton Down, UK. The HAI was developed at Public Health England laboratory and is based on the World Health Organization recommendations and is fit for purpose. Briefly, HAI was performed with human sera treated with receptor destroying enzyme incubated with standardized concentrations (4 haemagglutinin units) of influenza virus representing H1N1, H3N2 and influenza B (Victoria and Yamagata) 2020/21 viral strains. Chicken red blood cells were utilised as the source of RBCs for haemagglutination. HAI titres were determined as the reciprocal of the highest dilution of serum that completed inhibited haemagglutination.

*SARS-CoV-2 spike protein serum IgG ELISA (performed at Novavax Clinical Immunology Laboratory, Gaithersburg, MD, USA)*

Recombinant SARS-CoV-2 (rSARS-CoV-2) S protein was immobilised onto the surface of the 96-well microtitre plate wells (100 µL per well) by direct adsorption for 15 to 72 hours at 2°C to 8°C at a concentration of 1 µg/mL in PBS as per P_SOP_02483 (Validated method). Plates were washed 4 times with 300 µL/well PBST, blocked with 300 µL blocking buffer for 1-1.5 hours at 24°C ± 2°C. Diluted reference standard (2-fold dilution series of 12 dilutions starting 1:1000) and human serum samples (3-fold dilution series of 12 dilutions) in assay buffer (1% milk in PBS) starting at 1:100 dilution are then added in duplicate (100 µL per well) to the rSARS-CoV-2 S protein-coated wells and any specific antibodies are allowed to complex with the coated antigen for 2 hours ± 10 minutes at 24°C ± 2°C. Plates are washed 4 times with 300 µL/well PBST. Antibodies bound to the rSARS-CoV-2 S protein are then detected using a horseradish peroxidase (HRP) conjugate goat anti-human IgG antibody diluted 1: 2000 (Southern Biotech cat no. 2040-05) incubated for 1 hour ± 10 minutes at 24°C ± 2°C, washed 6 times with 300 µL/well PBST, and a colorimetric signal generated by addition of 100 µL per well 3, 3′,5,5′-tetramethylbenzidine (TMB) chromogenic substrate for 10 minutes ± 2 minutes at 24°C ± 2°C. After incubation was complete, the TMB reaction was stopped with 100 µL per well of TMB Stop solution. The absorbance was measured at 450 nm using a Molecular Device 96-well plate reader. When binding reagents (coated antigen and secondary antibody) are in excess, the optical density (OD) of the chromogenic substrate at endpoint is proportional to the quantity of anti-rSARS-CoV-2 S IgG present in the serum sample. The total anti-rSARS-CoV-2 S protein IgG antibody level in a serum sample was quantitated in ELISA unit, EU/mL, by comparison to a reference standard curve. The results were analysed in singleton by SoftMax Pro software using 4-PL curve fit. Assay included control plates comprising of positive controls and negative control.

**Supplemental Tables**

**Table S1. Potential Immune-Mediated Medical Conditions (PIMMC)**

| **Categories** | **Diagnoses (as MedDRA Preferred Terms)** |
| --- | --- |
| Neuro-inflammatory Disorders: | Acute disseminated encephalomyelitis (including site specific variants: eg, non-infectious encephalitis, encephalomyelitis, myelitis, myeloradiculomyelitis), cranial nerve disorders including paralyses/paresis (eg, Bell’s palsy), generalised convulsion, Guillain-Barre syndrome (including Miller Fisher syndrome and other variants), immune‑mediated peripheral neuropathies and plexopathies (including chronic inflammatory demyelinating polyneuropathy, multifocal motor neuropathy and polyneuropathies associated with monoclonal gammopathy), myasthenia gravis, multiple sclerosis, narcolepsy, optic neuritis, transverse myelitis, uveitis |
| Musculoskeletal and Connective Tissue Disorders: | Anti-synthetase syndrome, dermatomyositis, juvenile chronic arthritis (including Still’s disease), mixed connective tissue disorder, polymyalgia rheumatic, polymyositis, psoriatic arthropathy, relapsing polychondritis, rheumatoid arthritis, scleroderma (including diffuse systemic form and CREST syndrome), spondyloarthritis (including ankylosing spondylitis, reactive arthritis [Reiter's syndrome] and undifferentiated spondyloarthritis), systemic lupus erythematosus, systemic sclerosis, Sjogren’s syndrome |
| Vasculitides: | Large vessels vasculitis (including giant cell arteritis such as Takayasu's arteritis and temporal arteritis), medium sized and/or small vessels vasculitis (including polyarteritis nodosa, Kawasaki's disease, microscopic polyangiitis, Wegener's granulomatosis, Churg–Strauss syndrome [allergic granulomatous angiitis], Buerger’s disease [thromboangiitis obliterans], necrotising vasculitis and anti-neutrophil cytoplasmic antibody [ANCA] positive vasculitis [type unspecified], Henoch-Schonlein purpura, Behcet's syndrome, leukocytoclastic vasculitis) |
| Gastrointestinal Disorders: | Crohn’s disease, celiac disease, ulcerative colitis, ulcerative proctitis |
| Hepatic Disorders: | Autoimmune hepatitis, autoimmune cholangitis, primary sclerosing cholangitis, primary biliary cirrhosis |
| Renal Disorders: | Autoimmune glomerulonephritis (including IgA nephropathy, glomerulonephritis rapidly progressive, membranous glomerulonephritis, membranoproliferative glomerulonephritis, and mesangioproliferative glomerulonephritis. |
| Cardiac Disorders: | Autoimmune myocarditis/cardiomyopathy |
| Skin Disorders: | Alopecia areata, psoriasis, vitiligo, Raynaud’s phenomenon, erythema nodosum, autoimmune bullous skin diseases (including pemphigus, pemphigoid and dermatitis herpetiformis), cutaneous lupus erythematosus, morphoea, lichen planus, Stevens-Johnson syndrome, Sweet’s syndrome |
| Haematologic Disorders: | Autoimmune haemolytic anaemia, autoimmune thrombocytopenia, antiphospholipid syndrome, thrombocytopenia |
| Metabolic Disorders: | Autoimmune thyroiditis, Grave’s or Basedow’s disease, Hashimoto thyroiditis, diabetes mellitus type 1, Addison’s disease |
| Other Disorders: | Goodpasture syndrome, idiopathic pulmonary fibrosis, pernicious anaemia, sarcoidosis |
| Abbreviations: ANCA=anti-neutrophil cytoplasmic antibody; IgA=immunoglobulin A; MedDRA=*Medical Dictionary for Regulatory Activities.* | |

**Table S2. Adverse Events of Special Interest Relevant to COVID-19^*^**

| **Body System** | **Diagnoses** |
| --- | --- |
| Immunologic | Enhanced disease following immunisation, cytokine release syndrome related to COVID-19, multisystem inflammatory syndrome in children (MIS-C) |
| Respiratory | Acute respiratory distress syndrome (ARDS) |
| Cardiac | Acute cardiac injury including:   - Microangiopathy - Heart failure and cardiogenic shock - Stress cardiomyopathy - Coronary artery disease - Arrhythmia - Myocarditis, pericarditis |
| Haematologic | Coagulation disorder   - Deep vein thrombosis - Pulmonary embolus - Cerebrovascular stroke - Limb ischaemia - Haemorrhagic disease - Thrombotic complications |
| Renal | Acute kidney injury |
| Gastrointestinal | Liver injury |
| Neurologic | Guillain-Barré Syndrome, anosmia, ageusia, meningoencephalitis |
| Dermatologic | Chilblain-like lesions, single organ cutaneous vasculitis, erythema multiforme |
| Abbreviations: AESI=adverse event of special interest; COVID-19=coronavirus disease 2019; DAIDS=Division of AIDS; PCR=polymerase chain reaction; SARS-CoV2=severe acute respiratory syndrome coronavirus 2.  *To be recorded as AESIs relevant to COVID-19, these complications should be associated with a positive PCR test for SARS-CoV-2. | |

**Table S3. FDA Toxicity Grading Scale for Clinical Abnormalities (Local and General Systemic Reactogenicity)**

| **Local Reaction to Injectable Product** | | | | |
| --- | --- | --- | --- | --- |
| **Clinical Abnormality** | **Mild (Grade 1)** | **Moderate**  **(Grade 2)** | **Severe (Grade 3)** | **Potentially Life-Threatening (Grade 4)** |
| Pain | Does not interfere with activity | Repeated use of non-narcotic pain reliever >24 hours or interferes with activity | Any use of narcotic pain reliever or prevents daily activity | ER visit or hospitalisation |
| Tenderness | Mild discomfort to touch | Discomfort with movement | Significant discomfort at rest | ER visit or hospitalisation |
| Erythema/redness^a^ | 2.5−5 cm | 5.1−10 cm | >10 cm | Necrosis or exfoliative dermatitis |
| Induration/swelling^b^ | 2.5−5 cm and does not interfere with activity | 5.1−10 cm or interferes with activity | >10 cm or prevents daily activity | Necrosis |
| **Systemic (General)** | | | | |
| **Clinical Abnormality** | **Mild (Grade 1)** | **Moderate**  **(Grade 2)** | **Severe (Grade 3)** | **Potentially Life-Threatening (Grade 4)** |
| Fever (°C)  (°F) | 38.0−38.4 100.4−101.1 | 38.5−38.9 101. −102.0 | 39.0−40.0 102.1−104.0 | >40.0 >104.0 |
| Nausea/vomiting | No interference with activity or 1−2 episodes/24 hours | Some interference with activity or >2 episodes/24 hours | Prevents daily activity, or requires outpatient IV hydration | ER visit or hospitalisation for hypotensive shock |
| Headache | No interference with activity | Repeated use of non-narcotic pain reliever >24 hours or some interference with activity | Significant; any use of narcotic pain reliever or prevents daily activity | ER visit or hospitalisation |
| Fatigue/Malaise | No interference with activity | Some interference with activity | Significant; prevents daily activity | ER visit or hospitalisation |
| Myalgia | No interference with activity | Some interference with activity | Significant; prevents daily activity | ER visit or hospitalisation |
| Arthralgia | No interference with activity | Some interference with activity | Significant; prevents daily activity | ER visit or hospitalisation |
| Abbreviations: DHHS=US Department of Health and Human Services; ER=emergency room; FDA=US Food and Drug Administration. | | | | |

| **Table S4. Demographics for the Influenza Vaccination Sub-Study Population, Reactogenicity Cohort, and Intention-to-Treat Group (Without the Influenza Sub-Study Participants)** | | | | | |
| --- | --- | --- | --- | --- | --- |
|  | **NVX-CoV2373 + Influenza**  **(n=217)** | **Placebo +**  **Influenza**  **(n=214)** | **Total Influenza**  **Sub-Study**  **(n=431)** | **Reactogenicity Cohort**  **(Excluding Influenza**  **Sub-Study)**  **(n=2310)** | **ITT Population**  **(Excluding Influenza**  **Sub-Study)**  **(n=14708)** |
| Age, yr (SD)  Median  Range | 42.3 (14.09)  40.0  20, 71 | 41.9 (13.23)  38.0  23, 77 | 42.1 (13.66)  39.0  20, 77 | 50.2 (15.07)  52.0  18, 84 | 53.5 (14.82)  56.0  18, 84 |
| Age group, n (%)  18-64 yr (SD)  ≥65 yr (SD) | 201 (92.6)  16 (7.3) | 201 (93.9)  13 (6.1) | 402 (93.3)  29 (6.7) | 1851 (80.1)  459 (19.9) | 10612 (72.2)  4096 (27.8) |
| Sex, n (%)  Male  Female | 123 (56.7)  94 (43.3) | 118 (55.1)  96 (44.9) | 241 (55.9)  190 (44.1) | 1105 (47.8)  1205 (52.2) | 7567 (51.4)  7141 (48.6) |
| Race or ethnic group, n (%)  White  Black or African American  Asian  Multiple  Not reported  Other  Missing  Hispanic or Latinx | 163 (75.1)  4 (1.8)  14 (6.5)  29 (13.3)  3 (1.4)  3 (1.4)  1 (0.4)  10 (4.6) | 164 (76.6)  2 (0.9)  23 (10.7)  23 (10.7)  2 (0.9)  0 (0)  0 (0)  5 (2.3) | 327 (75.9)  6 (1.4)  37 (8.6)  52 (12.1)  5 (1.2)  3 (0.7)  1 (0.2)  15 (3.5) | 2218 (96.0)  5 (0.2)  47 (2.0)  8 (1.3)  30 (1.3)  0 (0)  2 (<0.1)  12 (0.5) | 13953 (94.9)  54 (0.4)  425 (2.9)  84 (0.6)  171 (1.2)  14 (<0.1)  7 (<0.1)  110 (0.7) |
| Co-morbidity status  Yes  No | 55 (25.3)  162 (74.7) | 62 (29.0)  152 (71.0) | 117 (27.1)  314 (72.9) | 1032 (44.7)  1278 (55.3) | 6650 (45.2)  8054 (54.8) |
| Abbreviation: ITT=intention-to-treat; SD=standard deviation. | | | | | |

Abbreviations: IgG=immunoglobulin G; ITT=intention-to-treat; SARS-CoV-2=severe acute respiratory syndrome coronavirus 2; SD=standard deviation.

| **Table S5. Demographics for the Anti-S IgG Immunogenicity Cohort and Intention-to-Treat Population** | | | |
| --- | --- | --- | --- |
|  | **Immunogenicity Cohort, NVX-CoV2373**  **(n=502)** | **Immunogenicity Cohort, Placebo**  **(n=497)** | **Total, ITT Population**  **(n=15139)** |
| Age, yr (SD)  Median  Range | 51.6 (15.71)  54.0  18, 81 | 51.4 (15.39)  52  19, 93 | 53.1 (14.91)  55.0  18, 84 |
| Age group, n (%)  18-64 yr (SD)  ≥65 yr (SD) | 370 (73.7)  132 (26.3) | 368 (74.0)  129 (26.0) | 11014 (72.8)  4125 (27.2) |
| Sex, n (%)  Male  Female | 258 (51.4)  244 (48.6) | 290 (58.4)  207 (41.6) | 7808 (51.6)  7331 (48.4) |
| Race or ethnic group, n (%)  White  Black or African American  Asian  Multiple  Not reported  Other  Missing  Hispanic or Latinx | 437 (87.1)  3 (0.6)  39 (7.8)  4 (0.8)  18 (3.6)  1 (0.2)  0  10 (2.0) | 436 (87.7)  3 (0.6)  34 (6.8)  2 (0.4)  21 (4.2)  1 (0.2)  0  6 (1.2) | 14280 (94.3)  60 (0.4)  462 (3.1)  136 (0.9)  176 (1.2)  17 (<0.1)  8  125 (0.8) |
| SARS-CoV-2 serostatus,  n (%)  Negative  Positive  Missing | 475 (94.6)  24 (3.8)  3 | 475 (95.6)  20 (4.0)  2 | 14362 (94.9)  643 (4.2)  134 |
| Co-morbidity status  Yes  No | 171 (34.1)  331 (65.9) | 170 (34.2)  327 (65.8) | 6767 (44.7)  8372 (55.3) |

**Table S6. Reactogenicity for Participants in the Influenza Vaccination Sub-Study and Reactogenicity Cohort After Dose 1, All Ages**

|  | **NVX-CoV2373 + Influenza Vaccine (n=205)** | | | | | **NVX-CoV2373 Alone (n=1159)** | | | | | **Placebo + Influenza Vaccine (n=199)** | | | | | **Placebo Alone (n=1151)** | | | | |
| --- | --- | --- | --- | --- | --- | --- | --- | --- | --- | --- | --- | --- | --- | --- | --- | --- | --- | --- | --- | --- |
|  | **Grade 0** | **Grade 1** | **Grade 2** | **Grade 3** | **Grade 4** | **Grade 0** | **Grade 1** | **Grade 2** | **Grade 3** | **Grade 4** | **Grade 0** | **Grade 1** | **Grade 2** | **Grade 3** | **Grade 4** | **Grade 0** | **Grade 1** | **Grade 2** | **Grade 3** | **Grade 4** |
| **A) Local AEs** |  |  |  |  |  |  |  |  |  |  |  |  |  |  |  |  |  |  |  |  |
| Any local AE | 29.9 | 52.3 | 16.1 | 1.7 | 0.0 | 42.4 | 41.5 | 15.1 | 1.0 | 0.0 | 60.6 | 27.2 | 12.2 | 0.0 | 0.0 | 82.1 | 14.7 | 3.0 | 0.2 | 0.0 |
| Pain | 60.3 | 35.1 | 4.0 | 0.6 | 0.0 | 70.7 | 28.4 | 0.9 | 0.0 | 0.0 | 83.3 | 16.1 | 0.6 | 0.0 | 0.0 | 90.8 | 9.0 | 0.1 | 0.1 | 0.0 |
| Tenderness | 35.1 | 48.3 | 14.9 | 1.7 | 0.0 | 46.7 | 37.8 | 14.5 | 1.0 | 0.0 | 62.8 | 25.0 | 12.2 | 0.0 | 0.0 | 85.7 | 11.4 | 2.7 | 0.1 | 0.0 |
| Erythema | 98.9 | 1.1 | 0.0 | 0.0 | 0.0 | 97.9 | 1.8 | 0.3 | 0.0 | 0.0 | 98.9 | 1.1 | 0.0 | 0.0 | 0.0 | 99.7 | 0.3 | 0.0 | 0.0 | 0.0 |
| Swelling | 98.9 | 0.0 | 1.1 | 0.0 | 0.0 | 99.1 | 0.6 | 0.3 | 0.0 | 0.0 | 100.0 | 0.0 | 0.0 | 0.0 | 0.0 | 99.5 | 0.4 | 0.2 | 0.0 | 0.0 |
| **B) Systemic AEs** |  |  |  |  |  |  |  |  |  |  |  |  |  |  |  |  |  |  |  |  |
| Any systemic AE | 39.9 | 37.0 | 20.2 | 2.9 | 0.0 | 54.3 | 28.7 | 15.7 | 1.1 | 0.2 | 52.8 | 23.3 | 21.1 | 2.8 | 0.0 | 63.7 | 22.4 | 12.8 | 1.1 | 0.0 |
| Nausea/vomiting | 94.8 | 4.6 | 0.6 | 0.0 | 0.0 | 94.8 | 4.3 | 0.8 | 0.0 | 0.1 | 93.3 | 5.6 | 1.1 | 0.0 | 0.0 | 94.8 | 3.8 | 1.4 | 0.0 | 0.0 |
| Headache | 75.1 | 17.9 | 6.4 | 0.6 | 0.0 | 75.5 | 20.0 | 4.0 | 0.5 | 0.1 | 78.3 | 15.6 | 6.1 | 0.0 | 0.0 | 78.5 | 16.7 | 4.6 | 0.3 | 0.0 |
| Fatigue | 72.3 | 13.3 | 13.3 | 1.2 | 0.0 | 80.6 | 10.3 | 8.7 | 0.4 | 0.1 | 71.1 | 11.7 | 15.6 | 1.7 | 0.0 | 82.4 | 9.1 | 8.2 | 0.3 | 0.0 |
| Malaise | 85.5 | 7.5 | 5.8 | 1.2 | 0.0 | 88.8 | 6.4 | 4.5 | 0.2 | 0.1 | 83.3 | 10.0 | 5.6 | 1.1 | 0.0 | 91.6 | 4.0 | 4.2 | 0.2 | 0.0 |
| Muscle pain | 71.7 | 19.7 | 8.7 | 0.0 | 0.0 | 78.6 | 16.2 | 5.0 | 0.1 | 0.1 | 80.0 | 14.4 | 5.6 | 0.0 | 0.0 | 86.7 | 10.3 | 2.6 | 0.4 | 0.0 |
| Joint pain | 92.5 | 6.4 | 1.2 | 0.0 | 0.0 | 93.6 | 4.4 | 1.9 | 0.0 | 0.1 | 98.3 | 1.7 | 0.0 | 0.0 | 0.0 | 94.5 | 3.0 | 2.3 | 0.2 | 0.0 |
| Temperature | 95.7 | 2.5 | 1.2 | 0.6 | 0.0 | 98.0 | 0.4 | 1.1 | 0.4 | 0.1 | 98.3 | 0.6 | 0.0 | 1.2 | 0.0 | 98.5 | 1.0 | 0.5 | 0.0 | 0.0 |
| Abbreviation: AE=adverse event. | | | | | | | | | | | | | | | | | | | | |

**Table S7. Reactogenicity for Participants in the Influenza Vaccination Sub-Study and Reactogenicity Cohort After Dose 1, Participants 18 to <65 Years Old**

|  | **NVX-CoV2373 + Influenza Vaccine (n=190)** | | | | | | **NVX-CoV2373 Alone (n=931)** | | | | | | **Placebo + Influenza Vaccine (n=186)** | | | | | | **Placebo Alone (n=920)** | | | | | |
| --- | --- | --- | --- | --- | --- | --- | --- | --- | --- | --- | --- | --- | --- | --- | --- | --- | --- | --- | --- | --- | --- | --- | --- | --- |
|  | **Any Gr** | **Gr 0** | **Gr 1** | **Gr 2** | **Gr 3** | **Gr 4** | **Any Gr** | **Gr 0** | **Gr 1** | **Gr 2** | **Gr 3** | **Gr 4** | **Any Gr** | **Gr 0** | **Gr 1** | **Gr 2** | **Gr 3** | **Gr 4** | **Any Gr** | **Gr 0** | **Gr 1** | **Gr 2** | **Gr 3** | **Gr 4** |
| **A) Local AEs** |  |  |  |  |  |  |  |  |  |  |  |  |  |  |  |  |  |  |  |  |  |  |  |  |
| Any local AE | 72.7 | 27.3 | 53.4 | 17.4 | 1.9 | 0.0 | 63.0 | 37.0 | 44.0 | 17.7 | 1.2 | 0.0 | 39.1 | 60.9 | 26.0 | 13.0 | 0.0 | 0.0 | 20.5 | 79.5 | 16.8 | 3.5 | 0.2 | 0.0 |
| Pain | 41.6 | 58.4 | 36.6 | 4.3 | 0.6 | 0.0 | 33.3 | 66.7 | 32.1 | 1.1 | 0.0 | 0.0 | 15.4 | 84.6 | 14.8 | 0.6 | 0.0 | 0.0 | 10.6 | 89.4 | 10.4 | 0.1 | 0.1 | 0.0 |
| Tenderness | 67.1 | 32.9 | 49.1 | 16.1 | 1.9 | 0.0 | 57.7 | 42.3 | 39.6 | 16.9 | 1.2 | 0.0 | 37.3 | 62.7 | 24.3 | 13.0 | 0.0 | 0.0 | 16.6 | 83.4 | 13.4 | 3.1 | 0.1 | 0.0 |
| Erythema | 1.2 | 98.8 | 1.2 | 0.0 | 0.0 | 0.0 | 2.2 | 97.8 | 1.9 | 0.3 | 0.0 | 0.0 | 1.2 | 98.8 | 1.2 | 0.0 | 0.0 | 0.0 | 0.3 | 99.7 | 0.3 | 0.0 | 0.0 | 0.0 |
| Swelling | 1.2 | 98.8 | 0.0 | 1.2 | 0.0 | 0.0 | 1.1 | 98.9 | 0.8 | 0.3 | 0.0 | 0.0 | 0.0 | 100.0 | 0.0 | 0.0 | 0.0 | 0.0 | 0.7 | 99.3 | 0.5 | 0.2 | 0.0 | 0.0 |
| **B) Systemic AEs** |  |  |  |  |  |  |  |  |  |  |  |  |  |  |  |  |  |  |  |  |  |  |  |  |
| Any systemic AE | 61.9 | 38.1 | 36.9 | 21.9 | 3.1 | 0.0 | 49.8 | 50.2 | 31.0 | 17.3 | 1.2 | 0.2 | 46.7 | 53.3 | 23.1 | 21.9 | 1.8 | 0.0 | 39.6 | 60.4 | 23.6 | 14.7 | 1.3 | 0.0 |
| Nausea/  vomiting | 5.6 | 94.4 | 5.0 | 0.6 | 0.0 | 0.0 | 5.2 | 94.8 | 4.2 | 0.9 | 0.0 | 0.1 | 6.5 | 93.5 | 5.3 | 1.2 | 0.0 | 0.0 | 6.3 | 93.7 | 4.8 | 1.5 | 0.0 | 0.0 |
| Headache | 25.0 | 75.0 | 17.5 | 6.9 | 0.6 | 0.0 | 26.6 | 73.4 | 21.3 | 4.7 | 0.4 | 0.0 | 21.9 | 78.1 | 16.0 | 5.9 | 0.0 | 0.0 | 24.1 | 75.9 | 18.1 | 5.6 | 0.3 | 0.0 |
| Fatigue | 29.4 | 70.6 | 13.8 | 14.4 | 1.3 | 0.0 | 21.9 | 78.1 | 11.6 | 9.7 | 0.4 | 0.1 | 29.0 | 71.0 | 11.8 | 16.0 | 1.2 | 0.0 | 20.0 | 80.0 | 10.0 | 9.8 | 0.2 | 0.0 |
| Malaise | 15.6 | 84.4 | 8.1 | 6.3 | 1.3 | 0.0 | 12.1 | 87.9 | 7.0 | 4.7 | 0.2 | 0.1 | 16.6 | 83.4 | 10.1 | 5.9 | 0.6 | 0.0 | 9.8 | 90.2 | 4.7 | 4.8 | 0.2 | 0.0 |
| Muscle pain | 30.0 | 70.0 | 20.6 | 9.4 | 0.0 | 0.0 | 23.7 | 76.3 | 17.7 | 5.7 | 0.1 | 0.1 | 20.7 | 79.3 | 15.4 | 5.3 | 0.0 | 0.0 | 15.0 | 85.0 | 11.5 | 3.0 | 0.5 | 0.0 |
| Joint pain | 8.1 | 91.9 | 6.9 | 1.3 | 0.0 | 0.0 | 6.6 | 93.4 | 4.7 | 1.8 | 0.0 | 0.1 | 1.8 | 98.2 | 1.8 | 0.0 | 0.0 | 0.0 | 5.8 | 94.2 | 3.1 | 2.5 | 0.1 | 0.0 |
| Temperature | 4.0 | 96.0 | 2.0 | 1.3 | 0.7 | 0.0 | 2.2 | 97.8 | 0.5 | 1.2 | 0.5 | 0.1 | 0.6 | 99.4 | 0.0 | 0.0 | 0.6 | 0.0 | 1.4 | 98.6 | 1.1 | 0.4 | 0.0 | 0.0 |

Abbreviations: AE=adverse event; Gr=grade.

**Table S8. Reactogenicity for Participants in the Influenza Vaccination Sub-Study and Reactogenicity Cohort After Dose 1, Participants ≥65 Years Old**

|  | **NVX-CoV2373 + Influenza Vaccine (n=15)** | | | | | | **NVX-CoV2373 Alone (n=228)** | | | | | | **Placebo + Influenza Vaccine (n=13)** | | | | | | **Placebo Alone (n=231)** | | | | | |
| --- | --- | --- | --- | --- | --- | --- | --- | --- | --- | --- | --- | --- | --- | --- | --- | --- | --- | --- | --- | --- | --- | --- | --- | --- |
|  | **Any Gr** | **Gr 0** | **Gr 1** | **Gr 2** | **Gr 3** | **Gr 4** | **Any Gr** | **Gr 0** | **Gr 1** | **Gr 2** | **Gr 3** | **Gr 4** | **Any Gr** | **Gr 0** | **Gr 1** | **Gr 2** | **Gr 3** | **Gr 4** | **Any Gr** | **Gr 0** | **Gr 1** | **Gr 2** | **Gr 3** | **Gr 4** |
| **A) Local AEs** |  |  |  |  |  |  |  |  |  |  |  |  |  |  |  |  |  |  |  |  |  |  |  |  |
| Any local AE | 38.5 | 61.5 | 38.5 | 0.0 | 0.0 | 0.0 | 34.9 | 65.1 | 30.7 | 4.2 | 0.0 | 0.0 | 45.5 | 54.5 | 45.5 | 0.0 | 0.0 | 0.0 | 7.6 | 92.4 | 6.3 | 1.3 | 0.0 | 0.0 |
| Pain | 15.4 | 84.6 | 15.4 | 0.0 | 0.0 | 0.0 | 12.3 | 87.7 | 12.3 | 0.0 | 0.0 | 0.0 | 36.4 | 63.6 | 36.4 | 0.0 | 0.0 | 0.0 | 3.6 | 96.4 | 3.6 | 0.0 | 0.0 | 0.0 |
| Tenderness | 38.5 | 61.5 | 38.5 | 0.0 | 0.0 | 0.0 | 34.4 | 65.6 | 30.2 | 4.2 | 0.0 | 0.0 | 36.4 | 63.6 | 36.4 | 0.0 | 0.0 | 0.0 | 5.4 | 94.6 | 4.0 | 1.3 | 0.0 | 0.0 |
| Erythema | 0.0 | 100.0 | 0.0 | 0.0 | 0.0 | 0.0 | 1.4 | 98.6 | 1.4 | 0.0 | 0.0 | 0.0 | 0.0 | 100.0 | 0.0 | 0.0 | 0.0 | 0.0 | 0.0 | 100.0 | 0.0 | 0.0 | 0.0 | 0.0 |
| Swelling | 0.0 | 100.0 | 0.0 | 0.0 | 0.0 | 0.0 | 0.0 | 100.0 | 0.0 | 0.0 | 0.0 | 0.0 | 0.0 | 100.0 | 0.0 | 0.0 | 0.0 | 0.0 | 0.0 | 100.0 | 0.0 | 0.0 | 0.0 | 0.0 |
| **B) Systemic AEs** |  |  |  |  |  |  |  |  |  |  |  |  |  |  |  |  |  |  |  |  |  |  |  |  |
| Any systemic AE | 38.5 | 61.5 | 38.5 | 0.0 | 0.0 | 0.0 | 28.3 | 71.7 | 18.9 | 9.0 | 0.5 | 0.0 | 54.5 | 45.5 | 27.3 | 9.1 | 18.2 | 0.0 | 23.6 | 76.4 | 17.8 | 5.3 | 0.4 | 0.0 |
| Nausea/  vomiting | 0.0 | 100.0 | 0.0 | 0.0 | 0.0 | 0.0 | 5.2 | 94.8 | 4.7 | 0.5 | 0.0 | 0.0 | 9.1 | 90.9 | 9.1 | 0.0 | 0.0 | 0.0 | 0.9 | 99.1 | 0.0 | 0.9 | 0.0 | 0.0 |
| Headache | 23.1 | 76.9 | 23.1 | 0.0 | 0.0 | 0.0 | 15.6 | 84.4 | 14.2 | 0.9 | 0.5 | 0.0 | 18.2 | 81.8 | 9.1 | 9.1 | 0.0 | 0.0 | 11.6 | 88.4 | 11.1 | 0.4 | 0.0 | 0.0 |
| Fatigue | 7.7 | 92.3 | 7.7 | 0.0 | 0.0 | 0.0 | 9.0 | 91.0 | 4.7 | 4.2 | 0.0 | 0.0 | 27.3 | 72.7 | 9.1 | 9.1 | 9.1 | 0.0 | 8.0 | 92.0 | 5.3 | 2.2 | 0.4 | 0.0 |
| Malaise | 0.0 | 100.0 | 0.0 | 0.0 | 0.0 | 0.0 | 7.5 | 92.5 | 3.8 | 3.8 | 0.0 | 0.0 | 18.2 | 81.8 | 9.1 | 0.0 | 9.1 | 0.0 | 3.1 | 96.9 | 1.3 | 1.8 | 0.0 | 0.0 |
| Muscle pain | 7.7 | 92.3 | 7.7 | 0.0 | 0.0 | 0.0 | 11.8 | 88.2 | 9.9 | 1.9 | 0.0 | 0.0 | 9.1 | 90.9 | 0.0 | 9.1 | 0.0 | 0.0 | 6.7 | 93.3 | 5.8 | 0.9 | 0.0 | 0.0 |
| Joint pain | 0.0 | 100.0 | 0.0 | 0.0 | 0.0 | 0.0 | 5.7 | 94.3 | 3.3 | 2.4 | 0.0 | 0.0 | 0.0 | 100.0 | 0.0 | 0.0 | 0.0 | 0.0 | 4.4 | 95.6 | 2.7 | 1.3 | 0.4 | 0.0 |
| Temperature | 8.3 | 91.7 | 8.3 | 0.0 | 0.0 | 0.0 | 1.0 | 99.0 | 0.0 | 1.0 | 0.0 | 0.0 | 20.0 | 80.0 | 10.0 | 0.0 | 10.0 | 0.0 | 1.8 | 98.2 | 0.9 | 0.9 | 0.0 | 0.0 |

Abbreviations: AE=adverse event; Gr=grade.

**Table S9. Reactogenicity for Participants in the Influenza Vaccination Sub-Study After Dose 2 (No Concomitant Influenza Vaccine), All Ages**

|  | **NVX-CoV2373 Alone (n=203)** | | | | | | **Placebo Alone (n=196)** | | | | | |
| --- | --- | --- | --- | --- | --- | --- | --- | --- | --- | --- | --- | --- |
|  | **Any Grade** | **Grade 0** | **Grade 1** | **Grade 2** | **Grade 3** | **Grade 4** | **Any Grade** | **Grade 0** | **Grade 1** | **Grade 2** | **Grade 3** | **Grade 4** |
| **A) Local** |  |  |  |  |  |  |  |  |  |  |  |  |
| Any local AE | 85.5 | 15.0 | 36.1 | 42.9 | 6.0 | 0.0 | 21.6 | 78.4 | 20.0 | 1.6 | 0.0 | 0.0 |
| Pain | 57.1 | 42.9 | 44.4 | 12.0 | 0.8 | 0.0 | 11.2 | 88.8 | 10.4 | 0.8 | 0.0 |  |
| Tenderness | 78.9 | 21.1 | 36.1 | 38.3 | 4.5 | 0.0 | 20.0 | 80.0 | 18.4 | 1.6 | 0.0 | 0.0 |
| Erythema | 11.3 | 88.7 | 3.8 | 4.5 | 3.0 | 0.0 | 0.0 | 100.0 | 0.0 | 0.0 | 0.0 | 0.0 |
| Swelling | 10.5 | 89.5 | 3.8 | 4.5 | 2.3 | 0.0 | 0.0 | 100.0 | 0.0 | 0.0 | 0.0 | 0.0 |
| **B) Systemic** |  |  |  |  |  |  |  |  |  |  |  |  |
| Any systemic AE | 69.7 | 30.3 | 25.0 | 37.1 | 6.8 | 0.8 | 37.9 | 62.1 | 14.5 | 20.2 | 3.2 | 0.0 |
| Nausea/vomiting | 10.6 | 89.4 | 8.3 | 2.3 | 0.0 | 0.0 | 4.8 | 95.2 | 3.2 | 1.6 | 0.0 | 0.0 |
| Headache | 46.2 | 53.8 | 25.8 | 19.7 | 0.8 | 0.0 | 18.5 | 81.5 | 12.1 | 5.6 | 0.8 | 0.0 |
| Fatigue | 46.2 | 53.8 | 11.4 | 32.6 | 2.3 | 0.0 | 25.8 | 74.2 | 6.5 | 16.9 | 2.4 | 0.0 |
| Malaise | 42.4 | 57.6 | 15.2 | 25.8 | 1.5 | 0.0 | 16.1 | 83.9 | 4.0 | 8.9 | 3.2 | 0.0 |
| Muscle pain | 47.0 | 53.0 | 25.0 | 19.7 | 2.3 | 0.0 | 11.3 | 88.7 | 5.6 | 5.6 | 0.0 | 0.0 |
| Joint pain | 17.4 | 82.6 | 6.1 | 10.6 | 0.8 | 0.0 | 4.0 | 96.0 | 2.4 | 1.6 | 0.0 | 0.0 |
| Temperature | 8.3 | 91.7 | 6.6 | 0.0 | 0.8 | 0.8 | 0.0 | 100.0 | 0.0 | 0.0 | 0.0 | 0.0 |

Abbreviation: AE=adverse event.

**Table S10. Influenza Haemagglutination Inhibition Assay: GMT, GMFR, and SCR Values by Influenza Strain for QIVc**

|  | **NVX-CoV2373 + QIVc**  **(n=201)** | | | | **Placebo + QIVc**  **(n=201)** | | | |
| --- | --- | --- | --- | --- | --- | --- | --- | --- |
|  | **Day 0** | | **Day 21** | | **Day 0** | | **Day 21** | |
|  |  | **(95% CI)** |  | **(95% CI)** |  | **(95% CI)** | **GMT** | **(95% CI)** |
| **GMT** |  |  |  |  |  |  |  |  |
| A/H1N1 | 11.0 | (8.9, 13.5) | 186.5 | (151.7, 229.3) | 12.9 | (10.5, 15.9) | 170.6 | (140.9, 206.6) |
| A/H3N2 | 34.0 | (27.6, 41.8) | 243.7 | (213.5, 278.1) | 37.3 | (30.0, 46.4) | 226.2 | (195.4, 262.7) |
| B/Victoria | 3.3 | (2.9, 3.7) | 9.7 | (7.9, 11.9) | 3.3 | (2.9, 3.7) | 9.4 | (7.8, 11.3) |
| B/Yamagata | 8.1 | (6.7, 9.8) | 38.8 | (32.2, 46.9) | 8.8 | (7.3, 10.6) | 39.0 | (33.2, 45.8) |
| **GMFR** |  |  |  |  |  |  |  |  |
| A/H1N1 |  |  | 17.0 | (13.4, 21.5) |  |  | 13.2 | (10.5, 16.5) |
| A/H3N2 |  |  | 7.2 | (5.8, 8.8) |  |  | 6.1 | (5.0, 7.4) |
| B/Victoria |  |  | 2.9 | (2.5, 3.5) |  |  | 2.9 | (2.4, 3.4) |
| B/Yamagata |  |  | 4.8 | (4.0, 5.7) |  |  | 4.4 | (3.8, 5.2) |
| **SCR** |  |  |  |  |  |  |  |  |
| A/H1N1 |  |  | 76.1 | (69.6, 81.9) |  |  | 71.5 | (64.6, 77.8) |
| A/H3N2 |  |  | 68.0 | (61.0, 74.5) |  |  | 62.2 | (54.9. 69.0) |
| B/Victoria |  |  | 14.2 | (9.7, 19.9) |  |  | 9.8 | (6.0, 14.9) |
| B/Yamagata |  |  | 32.5 | (26.0, 39.5) |  |  | 32.1 | (25.6, 39.2) |

Abbreviations: CI=confidence interval; GMFR=geometric mean fold rise; GMT= geometric mean titre; QIVc=influenza vaccine quadrivalent, cellular; SCR=seroconversion rate.

**Table S11. Influenza Haemagglutination Inhibition Assay: GMT, GMFR, and SCR Values by Influenza Strain for aTIV**

|  | **NVX-CoV2373 + aTIV**  **(n=16)** | | | | **Placebo + aTIV**  **(n=13)** | | | |
| --- | --- | --- | --- | --- | --- | --- | --- | --- |
|  | **Day 0** | | **Day 21** | | **Day 0** | | **Day 21** | |
|  |  | **(95% CI)** | **GMT** | **(95% CI)** | **GMT** | **(95% CI)** | **GMT** | **(95% CI)** |
| **GMT** |  |  |  |  |  |  |  |  |
| A/H1N1 | 23.6 | (9.9, 56.6) | 159.0 | (91.6, 275.9) | 5.8 | (2.9, 11.8) | 112.8 | (39.2, 325.1) |
| A/H3N2 | 41.5 | (16.0, 107.6) | 173.2 | (101.1, 297.1) | 28.2 | (12.0, 66.3) | 199.0 | (102.1, 387.8) |
| B/Victoria | 4.2 | (2.4, 7.3) | 11.8 | (5.9, 23.8) | 2.9 | (1.7, 5.1) | 21.9 | (8.6, 56.1) |
| B/Yamagata* | 8.4 | (3.9, 18.0) | 14.7 | (7.2, 29.9) | 7.5 | (4.1, 13.8) | 18.1 | (8.4, 39.4) |
| **GMFR** |  |  |  |  |  |  |  |  |
| A/H1N1 |  |  | 6.7 | (3.5, 13.1) |  |  | 19.3 | (8.0, 46.9) |
| A/H3N2 |  |  | 4.2 | (1.8, 9.9) |  |  | 7.1 | (2.7, 18.7) |
| B/Victoria |  |  | 2.8 | (1.5, 5.2) |  |  | 7.5 | (2.8, 20.2) |
| B/Yamagata* |  |  | 1.8 | (1.1, 2.8) |  |  | 2.4 | (1.4, 4.0) |
| **SCR** |  |  |  |  |  |  |  |  |
| A/H1N1 |  |  | 75.0 | (47.6, 92.7) |  |  | 72.7 | (39.0, 94.0) |
| A/H3N2 |  |  | 50.0 | (24.7, 75.3) |  |  | 54.5 | (23.4, 83.3) |
| B/Victoria |  |  | 12.5 | (1.6, 38.3) |  |  | 27.3 | (6.0, 61.0) |
| B/Yamagata* |  |  | 0.0 | (0.0, 20.6) |  |  | 9.1 | (0.2, 41.3) |

*Strain not contained in the vaccine.

Abbreviations: aTIV=adjuvanted trivalent influenza vaccine; CI=confidence interval; GMFR=geometric mean fold rise; GMT=geometric mean titre; SCR=seroconversion rate.

**Table S12a. Anti-S IgG on Day 0 and Day 35 in the Influenza Vaccination Sub-Study (a) and Immunogenicity Cohort (b) by Serostatus, in the ITT Population, All Ages**

|  |  | **NVX-CoV2373 + Influenza Vaccine** | | | |  | **Placebo + Influenza Vaccine** | | | |
| --- | --- | --- | --- | --- | --- | --- | --- | --- | --- | --- |
|  |  | **Day 0** | | **Day 35** | |  | **Day 0** | | **Day 35** | |
|  |  |  | **(95% CI)** |  | **(95% CI)** |  |  | **(95% CI)** |  | **(95% CI)** |
| **GMEU** |  |  |  |  |  |  |  |  |  |  |
| IIV + NVX-CoV2372 or placebo, all ages | n=214 | 146.5 | (128.1, 167.7) | 32724.4 | (27862.1, 38435.2) | n=214 | 129.8 | (116.8, 144.2) | 142.7 | (125.3, 162.6) |
| QIVc + NVX-CoV2373 or placebo, all ages, seronegative | n=198 | 117.9 | (109.1,127.5) | 30439.1 | (25713.4, 36033.5) | n=196 | 110.9 | (104.9, 117.3) | 121.9 | (110.0, 135.2) |
| aTIV+ NVX-CoV2373 or placebo, all ages, seropositive | n=19 | 1504.4 | (686.4, 3297.1) | 71115.6 | (46813.0, 108032.8) | n=13 | 1503.2 | (775.8, 2912.4) | 1624.4 | (1083.3, 2435.7) |
| **GMFR** |  |  |  |  |  |  |  |  |  |  |
| IIV + NVX-CoV2372 or placebo, all ages | n=214 |  |  | 223.2 | (183.9, 271.2) | n=214 |  |  | 1.1 | (1.0, 1.2) |
| QIVc + NVX-CoV2373 or placebo, all ages, seronegative | n=198 |  |  | 258.1 | (213.1, 312.5) | n=196 |  |  | 1.1 | (1.0, 1.2) |
| aTIV+ NVX-CoV2373 or placebo, all ages, seropositive | n=19 |  |  | 47.3 | (23.2, 96.0) | n=13 |  |  | 1.1 | (0.7, 1.8) |
| **SCR** |  |  |  |  |  |  |  |  |  |  |
| IIV + NVX-CoV2372 or placebo, all ages | n=214 |  |  | 97.6 | (94.6, 99.2) | n=214 |  |  | 3.0 | (1.1, 6.3) |
| QIVc + NVX-CoV2373 or placebo, all ages, seronegative | n=198 |  |  | 97.9 | (94.8, 99.4) | n=196 |  |  | 2.6 | (0.9, 6.1) |
| aTIV+ NVX-CoV2373 or placebo, all ages, seropositive | n=19 |  |  | 94.4 | (72.7, 99.9) | n=13 |  |  | 8.3 | (0.2, 38.5) |

Abbreviations: aTIV=adjuvanted trivalent influenza vaccine; GMEU=geometric mean ELISA unit; GMFR=geometric mean fold rise; ELISA=enzyme-linked immunosorbent assay; IgG=immunoglobulin G; IIV=inactivated influenza vaccine (both aTIV and QIVc); ITT=intention-to-treat; QIVc=influenza vaccine quadrivalent, cellular; S=spike; SCR=seroconversion rate.

**Table S12b.**

|  |  | **NVX-CoV2373 Alone** | | | |  | **Placebo Alone** | | | |
| --- | --- | --- | --- | --- | --- | --- | --- | --- | --- | --- |
|  |  | **Day 0** | | **Day 35** | |  | **Day 0** | | **Day 35** | |
|  |  |  | **(95% CI)** |  | **(95% CI)** |  |  | **(95% CI)** |  | **(95% CI)** |
| **GMEU** |  |  |  |  |  |  |  |  |  |  |
| NVX-CoV2373 or placebo alone, all ages | n=502 | 129.1 | (119.9, 138.9) | 46679.3 | (42206.2, 51626.4) | n=497 | 124.7 | (116.5, 133.5) | 129.5 | (119.5, 140.4) |
| NVX-CoV2373 or placebo alone, seronegative | n=475 | 112.2 | (107.6, 117.0) | 44229.9 | (39920.0, 49005.3) | n=475 | 110.8 | (106.6, 115.1) | 115.4 | (108.6, 122.6) |
| NVX-CoV2373 or placebo alone, seropositive | n=24 | 1698.8 | (994.8, 2900.9) | 125489.8 | (91186.3, 172697.9) | n=20 | 1771.7 | (915.0, 3430.2) | 1756.9 | (984.6, 3135.1) |
| **GMFR** |  |  |  |  |  |  |  |  |  |  |
| NVX-CoV2373 or placebo alone, all ages | n=502 |  |  | 361.6 | (324.6, 402.9) | n=497 |  |  | 1.0 | (1.0, 1.1) |
| NVX-CoV2373 or placebo alone, seronegative | n=475 |  |  | 394.3 | (354.8, 438.3) | n=475 |  |  | 1.0 | (1.0, 1.1) |
| NVX-CoV2373 or placebo alone, seropositive | n=24 |  |  | 73.9 | (46.8, 116.5) | n=20 |  |  | 1.0 | (0.8, 1.2) |
| **SCR** |  |  |  |  |  |  |  |  |  |  |
| NVX-CoV2373 or placebo alone, all ages | n=502 |  |  | 98.9 | (97.4, 99.6) | n=497 |  |  | 1.1 | (0.4, 2.6) |
| NVX-CoV2373 or placebo alone, seronegative | n=475 |  |  | 99.1 | (97.6, 99.7) | n=475 |  |  | 1.2 | (0.4, 2.7) |
| NVX-CoV2373 or placebo alone, seropositive | n=24 |  |  | 95.7 | (78.1, 99.9) | n=20 |  |  | 0.0 | (0.0, 17.6) |

Abbreviations: aTIV=adjuvanted trivalent influenza vaccine; GMEU=geometric mean ELISA unit; GMFR=geometric mean fold rise; ELISA=enzyme-linked immunosorbent assay; IgG=immunoglobulin G; IIV=inactivated influenza vaccine (both aTIV and QIVc); ITT=intention-to-treat; QIVc=influenza vaccine quadrivalent, cellular; S=spike; SCR=seroconversion rate.
